# Childhood temperamental, emotional, and behavioral predictors of clinical mood and anxiety disorders in adolescence

**DOI:** 10.1101/2022.03.14.22271392

**Authors:** Nora R. Bakken, Laurie J. Hannigan, Alexey Shadrin, Guy Hindley, Helga Ask, Ted Reichborn-Kjennerud, Martin Tesli, Ole A. Andreassen, Alexandra Havdahl

## Abstract

**Background:** Mood and anxiety disorders, often emerging during adolescence, account for a large share of the global burden of disability. Prospectively assessed premorbid early signs and trajectories can provide useful insights for early detection and development of these disorders.

**Methods:** Using the health registry linked Norwegian Mother, Father and Child Cohort Study (MoBa) of 110,367 children, we here examine cross-sectional and longitudinal association between temperamental traits, emotional and behavioral problems in childhood (0.5-8 years) and diagnosis of mood or anxiety (emotional) disorders in adolescence (10-18 years). We included birth year and sex, retrieved from the Medical Birth Registry of Norway, as covariates in all analyses.

**Results:** Logistic regression analyses showed consistent and increasing associations between childhood negative emotionality, behavioral and emotional problems and adolescent diagnosis of emotional disorders, present from 6 months of age (negative emotionality) and with similar magnitude of association for the associated traits. Latent profile analysis incorporating latent growth models identified five developmental profiles of emotional and behavioral problems. A profile of early increasing behavioral and emotional problems with combined symptoms at 8 years (1.3% of sample) was the profile most strongly associated with emotional disorders in adolescence (OR *vs*. reference: 5.00, 95% CI: 3.73-6.30).

**Conclusions:** We found a consistent and increasing association between negative emotionality, behavioral and emotional problems in early to middle childhood and mood and anxiety disorders in adolescence. A developmental profile coherent with early and increasing disruptive mood dysregulation across childhood was most predictive of adolescent emotional disorders. Our results highlight the importance of early emotional dysregulation and childhood as a formative period in the development of adolescent mood and anxiety disorders, supporting a potential for prevention and early intervention initiatives.

## INTRODUCTION

Mood and anxiety disorders are leading global causes of disability. According to the Global Burden of Disease study from 2019, 29 million and 47 million disability-adjusted life years (DALYs) were lost due to anxiety and depressive disorders respectively, accounting for the highest relative burden in children and adolescents (Vos et al., 2020). The disorders tend to present with a chronic and recurrent course, where co-occurrence with other mental disorders and continuity into adulthood is common (Beesdo-Baum & Knappe, 2012; Rohde, Lewinsohn, Klein, Seeley, & Gau, 2013). Early identification and treatment have been shown to significantly improve prognosis (Davey & Mcgorry, 2019), leading to calls for progress in prevention strategies for mood and anxiety disorders (Arango et al., 2018; Fusar-Poli et al., 2021).

Childhood and adolescence have been described as an optimal time for prevention and early intervention, targeting mental health problems in general, and anxiety and depression spesifically (Arango et al., 2018; Hoare, Callaly, & Berk, 2020). The period of childhood through adolescence is a risk phase for the first occurrence of symptoms of anxiety and depression (Beesdo-Baum & Knappe, 2012; Centers for Disease Control and Prevention, 2013). Furthermore, childhood and adolescence are critical periods for brain development (Fox, Levitt, & Nelson Iii, 2010), as well as psychosocial adaptations (Malik & Marwaha, 2022). However, detecting early stages of mood and anxiety disorders have proven difficult, especially in children and adolescents where symptoms tend to be nonspecific, developmentally dependent, and with greater individual variability (Creswell, Waite, & Cooper, 2014; Petito et al., 2020).

In order to develop prevention and early intervention efforts, better knowledge about the early signs and developmental trajectories associated with mood and anxiety disorders is needed (Arango et al., 2018). Longitudinal studies in child and adolescent populations suggest that children and adolescents differ in developmental trajectories of symptoms of depression (Shore, Toumbourou, Lewis, & Kremer, 2018), anxiety (Nandi, Beard, & Galea, 2009) and behavioral problems (Bongers, Koot, van der Ende, & Verhulst, 2004). However, there is a lack of knowledge about precursors to later mood and anxiety disorders that are observable in the earliest years of life (< 4 years) as well as the relative importance of temperamental traits and symptoms of mental disorders for later diagnostic outcomes (Nandi et al., 2009; Shore et al., 2018).

Here we leverage a prospective population-based pregnancy cohort, the Norwegian Mother, Father and Child Cohort Study (MoBa) of 114,326 children to examine when and how temperamental traits and emotional and behavioral problems in infancy to middle childhood (0.5 to 8 years of age) associate with mood and anxiety disorders in adolescence (10-18 years of age). We characterize trajectories of emotional and behavioral problems during childhood and assess their relationships to mood and anxiety disorders in adolescence.

## MATERIALS AND METHODS

### Study population

MoBa is a population-based pregnancy cohort study conducted by the Norwegian Institute of Public Health. Participants were recruited from all over Norway from 1999 to 2008. The women consented to participation in 41% of the pregnancies (Magnus et al., 2016). The current study is based on version 12 of the quality-assured data files released for research in January 2019. The study sample included all children in MoBa with data on at least one of the five assessment waves between age 6 months and 8 years and linkage data from the Norwegian Patient Registry (NPR) in adolescence (age 10-18 years) (n = 110,367). A participant flow chart is presented in supplementary Figure S1.

The establishment of the MoBa cohort and initial data collection was based on a license from the Norwegian Data Protection Agency and approval from The Regional Committees for Medical and Health Research Ethics, and the MoBa cohort is now following the regulations of the Norwegian Health Registry Act. The current study was approved by the administrative board of the Norwegian Mother, Father and Child Cohort Study led by the Norwegian Institute of Public Health and The Regional Committees for Medical and Health Research Ethics (2016/1226/REK sør-øst C) in Norway.

#### Temperament and personality

Infant temperament was measured at 6 months of age using questions from the Infant Characteristics Questionnaire (ICQ) (Bates, Freeland, & Lounsbury, 1979). Two dimensions were evaluated on a 7-point likert scale: 7 items from the fussy/difficult subscale (negative emotionality) and 2 items representing positive temperament. Early childhood temperament was measured by the four dimensions of Emotionality, Activity and Shyness Temperament Questionnaire (EAS) when the child was 18 months, 3 years and 5 years (Mathiesen & Tambs, 1999). Emotionality (negative emotionality), shyness, sociability and activity were measured by 3 items each. At 8 years, personality was evaluated by 6 items from each of the five dimensions of the Short Norwegian Hierarchical Personality Inventory for Children (NHiPIC-30): Neuroticism (negative emotionality), extraversion, benevolence (agreeableness), imagination and conscientiousness (Vollrath, Hampson, & Torgersen, 2016). Due to theoretical continuity (Buss & Plomin, 2015), overlapping phenotypes and high-correlation between fussy temperament (ICQ), emotionality (EAS) and neuroticism (NHiPIC-30) we have interpreted the measures as a common continuum of negative emotionality (See Figure S2 and S3).

#### Emotional and behavioral problems

The Child Behavior Checklist (CBCL) was used to measure emotional problems (internalizing items) and behavioral problems (externalizing items) at 18 months, 3 years and 5 years (Achenbach & Ruffle, 2000). More specific measures of symptoms of emotional and behavioral disorders were used at 8 years. A 13-item Short Mood and Feelings Questionnaire (SMFQ) measured depressive symptoms (Angold, Costello, Messer, & Pickles, 1995). Symptoms of anxiety were measured on a 5-item short form of Screen for Child Anxiety Related Disorders (SCARED) (Birmaher et al., 1999). Symptoms of conduct problems (8 items), oppositional defiant problems (8 items), hyperactivity (9 items) and inattention (9 items) were assessed by the Rating Scale for Disruptive Behaviour Disorders (RS-DBD) (Silva et al., 2005).

An overview of individual items included in each measure is presented in Table S1. Ordinal Cronbach alpha for each scale is presented in Table S2.

#### Emotional (mood or anxiety) disorder in adolescence (10-18 years of age)

Information on diagnoses in adolescence was retrieved from the Norwegian Patient Registry comprising International Classification of Diseases Tenth Revision (ICD-10) (World Health Organization, 1990) diagnoses registered in specialist health care services from 2008 through 2018. As anxiety and mood disorders commonly co-occur and have overlapping symptomatology, we examined the disorders as a combined category of emotional disorders optimizing utility for the purpose of prevention and early detection (Bullis, Boettcher, Sauer-Zavala, Farchione, & Barlow, 2019). **Emotional disorders** comprised mood disorders including depressive and bipolar disorders (F30-F39), anxiety disorders (F40-F41) and emotional disorders with onset usually occurring in childhood and adolescence (F92-F93) received from 10 years of age. We also defined non-overlapping subcategories of **depressive disorders** (F32-F34.1) and **anxiety disorders** (F40-F41 or F930, F931 or F932).

#### Covariates

Given known sex differences in the prevalence of emotional disorders and the wide range in birth years in MoBa, we included sex and birth year as covariates in all analyses. Information on participants’ sex and birth year were retrieved from the Medical Birth Registry of Norway (MBRN), a national health registry containing information about all births in Norway.

### Statistical analysis

Associations between measures of temperament/personality, emotional and behavioral problems in childhood (6 months-8 years) and diagnosis of emotional disorder in adolescence (10-18 years) were evaluated in separate logistic regression analyses. All measures were standardized with a mean of 0 and SD of 1 prior to analysis to improve comparability. To account for dependency in the data resulting from some mothers reporting on several children (siblings), all logistic regression analyses were calculated with Huber-White robust standard errors clustered by maternal identity (Long JA, 2020; Zeileis, 2004). We applied a 5 % alpha level and used False discovery rate (Benjamini-Hochberg) correction to account for multiple testing (Benjamini & Hochberg, 1995). Missing values were handled by case-wise deletion. To evaluate if our results were driven by mood or anxiety disorders, we conducted sensitivity analyses stratifying for the two main categories of emotional disorders.

Latent profile analysis incorporating latent growth models for longitudinally measured traits were used to evaluate how developmental trajectories of emotional and behavioral problems in childhood relate to diagnosis of emotional disorder in adolescence. Parallel process latent growth models were applied to the CBCL domains emotional and behavioral problems, then measures of more specific symptom domains from the 8-year questionnaires (SCARED, SMFQ, RS-DBD) were incorporated alongside the growth processes in latent profile analysis to classify the developmental profiles (for further details see (Hannigan et al., 2021) and figure S4). A manual 3-step maximum likelihood approach using Mplus was applied for profile generation and prediction of distal outcome (Nylund-Gibson, Grimm, & Masyn, 2019; Vermunt, 2010). Sibling relatedness was accounted for by clustering within mothers. Missing data were handled by full information maximum likelihood. Model selection was, in accordance with other developmental studies, determined by classical fit statistics, entropy, the substantive interpretability of each profile, and the proportion of individuals assigned to the smallest profile (we used a threshold of >1%) (Petersen, Qualter, & Humphrey, 2019).

R-version 4.0.3 was used for all analyses except parts of the developmental models utilizing Mplus version 8.3 (Muthén, 1998-2017). The R-package “Phenotools” (https://github.com/psychgen/phenotools), was used to calculate scores for the scales and prepare diagnostic data. For further details on data analyses see supplementary text 1.

#### Sensitivity analysis

To evaluate whether our results were affected by outcome definition, we repeated the analyses restricting the outcome to adolescent-onset emotional disorder (n = 2,878) and childhood-onset adolescent-persistent emotional disorder (n = 461) defined as first registered diagnoses after or before the age of 10 years respectively. Main analyses were also stratified for sex, assessing validity for both males and females.

#### Consideration of missing data and attrition

We examined whether response rates for childhood measures differed between children with and without adolescent diagnosis of emotional disorder (See Table S3). We also examined characteristics of individuals excluded from developmental profile analyses due to missing values in all relevant variables (n = 28,480) (See table S4).

## RESULTS

Demographic information and descriptive information on key study variables are presented in Table 1 and Table S5.

**Table 1.** Demographic information for the study sample

|  | **Emotional disorder**<br>**(n = 3,339)** | **No emotional disorder**<br>**(n = 107,028)** | **Total**<br>**(n = 110,367)** |
| --- | --- | --- | --- |
| Sex, male, n (%) | 1598 (47.86%) | 54912 (51.31%) | 56510 (51.20%) |
| Age end of follow-up, years,<br>*m* (SD) | 14.28 (2.11) | 13.00 (2.12) | 13.04 (2.13) |
| Age first emotional disorder,<br>years, *M*, (SD) | 12.13 (2.63) | - | 12.13 (2.63) |
| Highly educated mother or<br>father, n (%) | 1684 (61.37%) | 66406 (72.83%) | 68090 (72.49%) |
| Marital status, n (%)<sup>a</sup> | 3102 (92.90%) | 102355 (95.63%) | 105457 (95.55%) |
| Maternal age, *m*, (SD)<sup>b</sup> | 29.37 (4.92) | 30.14 (4.63) | 30.12 (4.64) |
| Parity, n (%) primiparous | 1516 (45.40%) | 46823 (43.75%) | 48339 (43.80%) |
| Mother's country of birth |  |  |  |
| Norway, n (%) | 3063 (93.56%) | 96519 (91.82%) | 99582 (91.87%) |
| Other high-income-<br>country, n (%) | 139 (4.25%) | 4798 (4.56%) | 4937 (4.55%) |
| Other GDB 7 super<br>region country, n (%) | 72 (2.20%) | 3804 (3.62%) | 3876 (3.58%) |
| **Co-occurring diagnoses** |  |  |  |
| OCD, n, (%) | 116 (3.47%) | 345 (0.32%) | 461 (0.42%) |
| ADHD, n, (%) | 613 (18.36%) | 3936 (3.68%) | 4549 (4.12%) |
| CD, n, (%) | 101 (3.02%) | 487 (0.46%) | 588 (0.53%) |

**Note:** **a:** n (%) married/registered partner/co-habitant. **b:** mean maternal age at pregnancy is calculated for
mothers between age 17 and 45 due to lack of information on age for mothers above or below this age-range (n =
58). Co-occurring diagnoses= additional diagnosis during follow-up. OCD= Obsessive-compulsive disorder.
ADHD= Attention-Deficit/Hyperactivity Disorder. CD= Conduct disorders.

Of the 110,367 included individuals, 3,339 were registered with a diagnosis of emotional disorder during follow-up, stratified into the following sub-categories: depressive disorder (n = 644), anxiety disorder (n = 1,423), depressive and anxiety disorder (n = 259) and other emotional disorder including bipolar disorder (n = 1,013).

### Individual childhood trait measure associations with adolescent emotional disorder

Results from logistic regression analysis indicated positive associations between emotional disorders in adolescence and traits of negative emotionality at 6 months, 18 months, 3 years, 5 years and 8 years (all p-values^FDR^ <0.001), as well as shyness at 5 years (p-value^FDR^ 0.001).

Emotional and behavioral problems measured at 18 months, 3 years, 5 years and 8 years of age were consistently and with increasing effect sizes associated with emotional disorder in adolescence (all p-values^FDR^ <0.001). Negative associations with emotional disorders were found for agreeableness, conscientiousness, imagination, and extraversion (all p-values^FDR^ <0.001). Results are presented in Figure 1 and Table S6.

**Figure 1.**
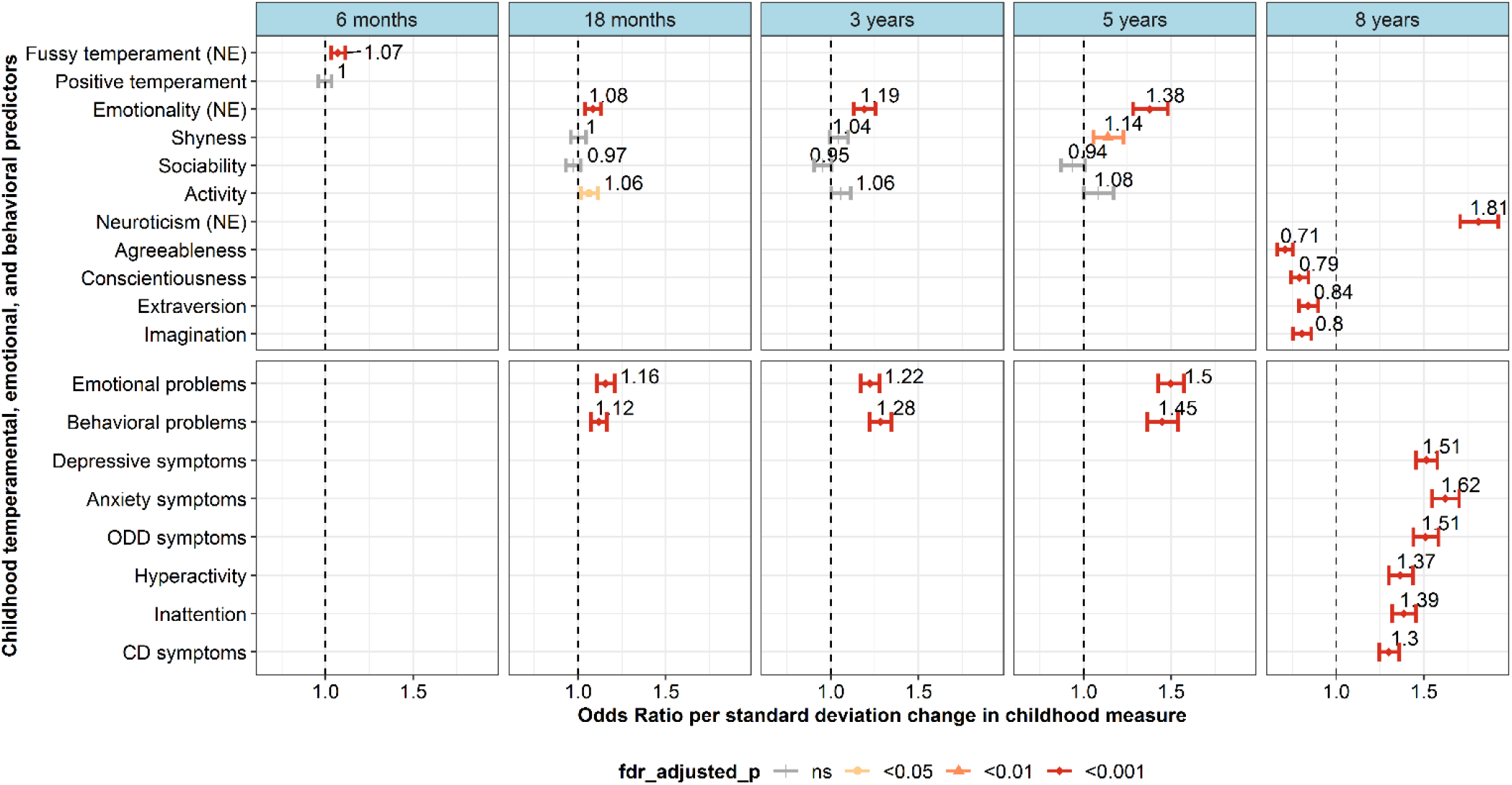
Association between temperamental traits and mental health problems in childhood and any emotional disorder in adolescence. **Note:** Odds ratio for the difference in odds of any emotional disorder in adolescence (10-18years) per standard deviation change in score for temperamental traits and mental health problems in childhood (6 months – 8 years). NE= Negative emotionality. Level of significance after false discovery rate (Benjamini-Hochberg) correction is illustrated by color and shape of point and error-bar.

Sensitivity analyses using the main subclasses of emotional disorders as outcomes (anxiety and depressive disorders) found similar association-patterns. Of notice, anxiety symptoms at 8 years were specifically associated with anxiety disorder and showed little association with depressive disorder in adolescence (10-18 years) (Figure 2 and Table S7). In contrast, depressive symptoms at 8 years were associated with all subclasses of emotional disorders.

**Figure 2:**
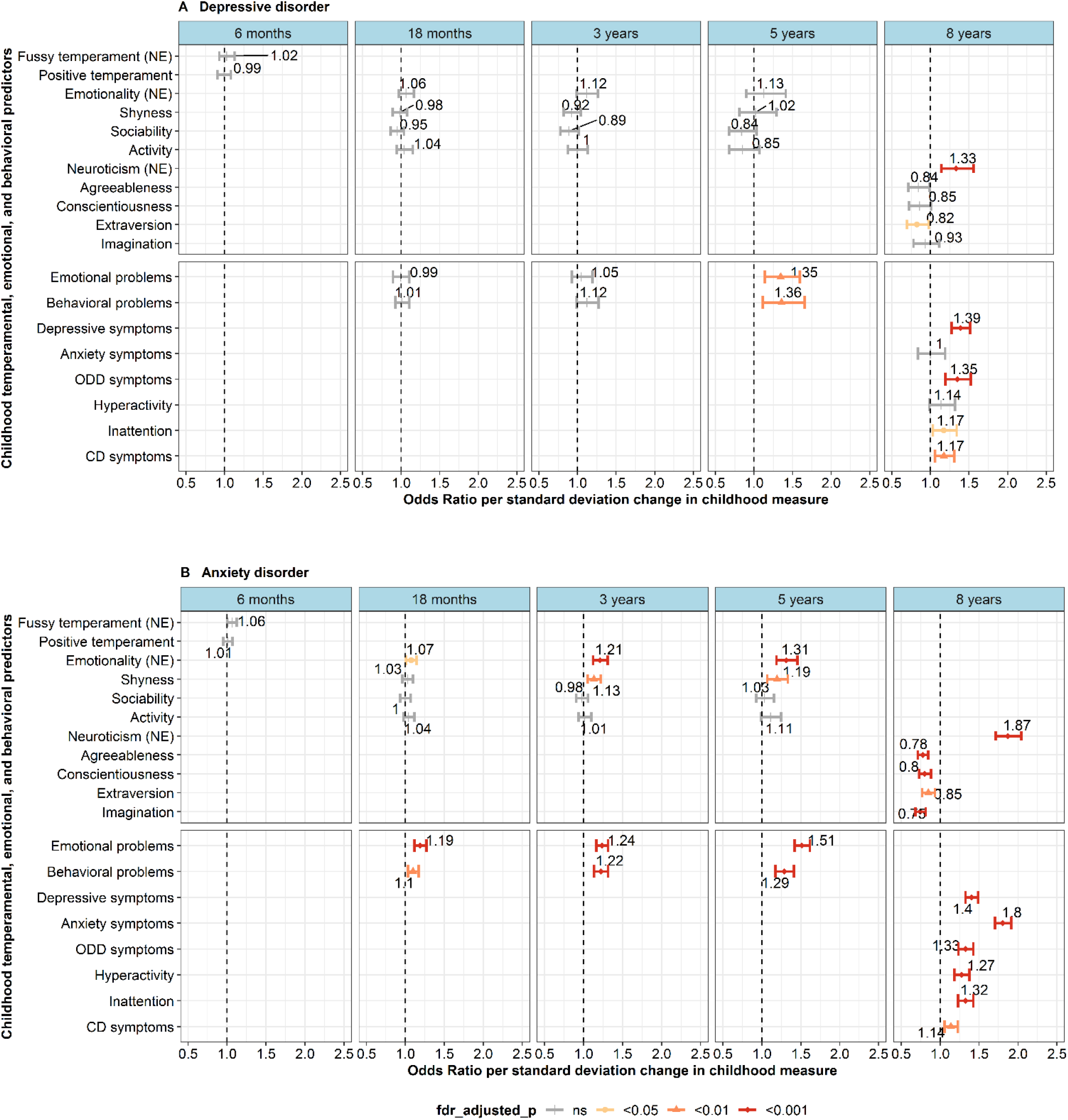
Association between temperamental traits and mental health problems in childhood and any anxiety or depressive disorder in adolescence. **Note:** Odds ratio for the difference in odds of depression(A) and anxiety(B) in adolescence (10-18 years) per standard deviation change in score for temperamental traits and mental health problems in childhood (6 months – 8 years). NE= Negative emotionality. Level of significance after false discovery rate (Benjamini-Hochberg) correction is illustrated by color and shape of point and error-bar. ns= not significant.

### Childhood developmental trajectories of emotional and behavioral problems as predictors of emotional disorders

Figure 3 and Table 2 summarize results for the disorder association to the developmental profiles of emotional and behavioral problems across childhood. We identified five distinct developmental profiles: **Profile 1 (3.8%):** Initially elevated and increasing behavioral problems followed by elevated levels of inattention and hyperactivity symptoms and moderate oppositional defiant disorder symptoms at 8 years. **Profile 2 (84.9%):** Initially low and decreasing early childhood emotional and behavioral problems and low symptoms at 8 years. **Profile 3 (4.9%):** Initially moderate and steeply increasing emotional problems followed by elevated anxiety symptoms at 8 years. **Profile 4 (1.3%):** Initially elevated and increasing behavioral problems along with initially low but increasing emotional problems in early childhood followed by elevated symptoms across a broad range of domains (depression, inattention, hyperactivity, oppositional defiant disorder and conduct disorder) at 8 years. **Profile 5 (5.1%):** Initially moderate and gradually decreasing early childhood behavioral problems with moderate levels of inattention, hyperactivity and oppositional defiant disorder symptoms at 8 years.

**Figure 3:**
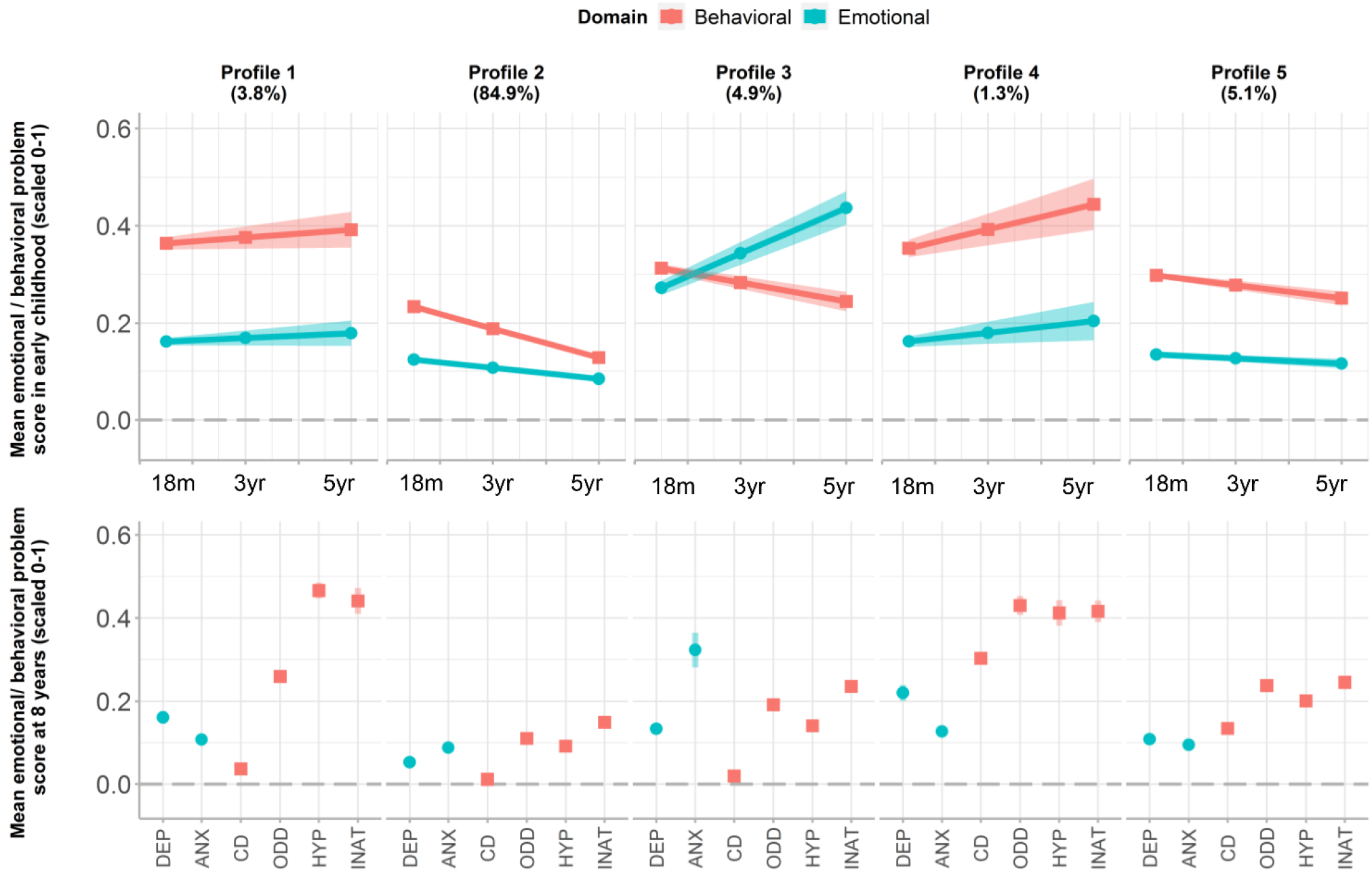
Developmental trajectories of childhood emotional and behavioral problems in early and middle childhood. **Note:** Figure illustrating developmental profiles of childhood emotional and behavioral problems. Upper panel shows development of early childhood emotional (blue, circle) and behavioral problems (red, square) measured by CBCL at 18 months, 3 years and 5 years of age. Lower panel shows corresponding middle childhood mental health problems. 95% confidence-intervals are illustrated in shaded colors. DEP= depressive symptoms. ANX= anxiety symptoms. CD= conduct disorder symptoms. ODD= oppositional defiant disorder problems. HYP= hyperactivity problems. INAT= inattention problems.

**Table 2.** Relative odds of any adolescent emotional disorder given assignment to specific developmental profile.

| Comparison | Odds ratio (OR) | CI |
| --- | --- | --- |
| Profile 1 vs Reference | 2.15 | (1.57, 2.74) |
| Profile 3 vs Reference | 3.49 | (2.88, 4.10) |
| Profile 4 vs Reference | 5.00 | (3.70, 6.30) |
| Profile 5 vs Reference | 1.37 | (1.05, 1.69) |
| Profile 1 vs profile 3 | 0.62 | (0.43, 0.81) |
| Profile 1 vs profile 4 | 0.43 | (0.27, 0.60) |
| Profile 1 vs profile 5 | 1.57 | (1.04, 2.11) |
| Profile 3 vs profile 4 | 0.70 | (0.49, 0.90) |
| Profile 3 vs profile 5 | 2.55 | (1.87, 3.22) |
| Profile 4 vs profile 5 | 3.65 | (2.47, 4.83) |

**Note:** Profile 2 including 84.9% (n = 69,522) of the individuals was used as reference class. Distribution of individuals to subsequent classes: Profile 1 (n = 3,087, 3.77%), Profile 3 (n = 4 045, 4.94%), Profile 4 (n = 1,074, 1.31%), Profile5 (n = 4,158, 5.01%). All profiles were compared to each other. All associations were significant with $p < 0.001$ .

Individuals classified in profile 1, 3 and 4, all characterized by increasing early childhood emotional and/or behavioral problems and elevated symptoms at 8 years, were all at least twice as likely to have an adolescent diagnosis of emotional disorder as individuals in the normative class with low symptoms throughout development (**Profile 1:** OR: 2.15, 95% CI: 1.57-2.74, **Profile 3:** OR: 3.49, 95% CI: 2.88-4.10, **Profile 4:** OR: 5.00, 95% CI: 3.70-6.30).

Fit statistics for assessed models and demographic characteristics including distribution of adolescent emotional diagnoses for the final five developmental profiles are presented in Table S8 and Table S9 respectively.

Sensitivity analyses evaluating implications of outcome definition and sex found overall the same pattern of associations in both logistic regression analyses and developmental analyses. Childhood-onset adolescent-persistent emotional disorders was generally stronger associated to the disorder-related phenotypes than the main outcome of adolescent-present emotional disorders (see TableS10 and Table S11). Similarly, some disorder-related traits (anxiety and depressive symptoms) and developmental-profiles (profile 4) were slightly weaker associated to adolescent-onset emotional disorders than adolescent present emotional disorders (see Table S12 and Table S13). Sensitivity analyses stratifying for sex identified the same pattern in individual symptom-relation and trajectories, however with profile 3 (dominated by emotional problems) being stronger associated with adolescent emotional disorders in males than in females (see Table S14, Table S15, Table S16 and Table S17).

## DISCUSSION

The main finding of the present study was that temperamental traits and emotional and behavioral problems in childhood are consistently and increasingly associated with diagnoses of mood and anxiety disorders in adolescence. We found consistent differences in mental traits for children later diagnosed with an emotional disorder from 6 months of age, expanding knowledge from previous studies of smaller sample size, and with shorter and/or later time of follow-up (Nandi et al., 2009; Shore et al., 2018). Distinct trajectories of emotional and behavioral problems in childhood related to later mood or anxiety disorders in adolescence were identified. Collectively, our findings emphasize the relevance of early childhood in development of mood and anxiety disorders. From a clinical perspective, we here show that integrated information of individuals’ emotional and behavioral functioning across childhood by the age of 8 years can help identify a subset of individuals with up to a 5-fold increased risk of mood and/or anxiety disorder in adolescence.

Our findings indicate that adolescent mood and anxiety disorders are often preceded by a gradual development of emotional and behavioral problems in early childhood. Expanding knowledge from previous studies, our analysis identified differences in mental traits present from 6 months of age, increasing in degree and specificity with aging. Coherent with previous studies, trajectories of increasing symptom-burden in childhood were predictive of adolescent diagnosis in our analyses (Battaglia et al., 2017; Dekker et al., 2007). Sensitivity analyses finding the strongest association to childhood-onset emotional disorders further supports mental traits potential as early signs or manifestations of disorder. Clinically our findings are of relevance for research on prevention and early identification efforts. As of today, there is no consensus regarding general screening initiatives aimed at children (Siu, 2016; Walter et al., 2020). Our study indicates that divergence from typical emotional and behavioral development can be detected already in childhood. However, the study also emphasizes the complexity and relatively small effect-sizes for signs and symptoms presenting in this age group, thus acknowledging the challenges related to identification of children with high-risk of adolescent disorder.

Emotional dysregulation stands out as an important characteristic in the development of mood and anxiety disorders. Negative emotionality, characterized by an increased propensity to experience and react with intense negative emotions, such as sadness, anxiety, fear and anger (Kann, O’Rawe, Huang, Klein, & Leung, 2017) was the temperamental trait consistently most strongly associated with later emotional disorder. Negative emotionality and emotional problems, both encompassed in the term of emotional dysregulation, are conceptualized as transdiagnostic traits related to the development and worse prognosis of mood and anxiety disorders (Dougherty et al., 2013). Association between emotional dysregulation and mood and anxiety disorders has been identified at the genetic and biological level, and has been found to be a predictor of future major depression and bipolar disorder in adults (Vidal-Ribas, Brotman, Valdivieso, Leibenluft, & Stringaris, 2016). Our study provides novel empirical support for the importance of emotional dysregulation in the development of mood and anxiety disorders, identifying a consistent and strengthening relationship from infancy to middle childhood.

The importance of emotional dysregulation is further emphasized by the relative importance of the developmental profile 4. This profile has characteristics coherent with the controversial disruptive mood dysregulation disorder (DMDD) included in the Diagnostic and Statistical Manual of Mental disorders (DSM-5), that is characterized by persistent irritability and recurrent temper outbursts (Rao, 2014). The combined phenotype characterizing this disorder and developmental profile, together with our findings of similar magnitude of association between emotional and behavioral problems and adolescent mood and anxiety disorders, highlight the role of dysregulation rather than emotional symptoms alone. Together this stresses the need to also consider behavioral problems as potential manifestations of emotional dysregulation or risk factors for emotional disorders, incorporating a broad developmental perspective when evaluating need for preventive interventions in this age group.

Results of our study must be interpreted while considering its limitations. Although the use of maternal reported prospective measures of childhood mental traits and clinician-assigned diagnostic outcome avoids common rating method bias, the maternal report can be influenced by characteristics of the mother. Additionally, clinician-assigned diagnosis is reliant on help-seeking and referral to specialist health care. Furthermore, some children in our analysis are young, thus additional diagnoses are expected in the following years. However, based on sensitivity-analysis finding similar pattern of associations for adolescence-present diagnosis (main outcome), adolescence-onset diagnosis and childhood-onset diagnosis, we expect our results to be robust to this limitation. Due to limited power, we did not test for sex-specific associations. However, because of unclear findings of sex differences in temperament, emotional and behavioral traits (Chaplin & Aldao, 2013), we conducted sex-stratified sensitivity analyses finding support for generalization of our results across sex. Despite population-based sampling methods, another limitation in our study is the possibility of reduced representativeness due to selection bias. The participation and attrition in the MoBa cohort in general has been thoroughly reported in other studies (Magnus et al., 2016; Magnus et al., 2006). Briefly, there is an underrepresentation of the youngest women (<25 years), mothers living alone, women with stillbirths and mothers smoking (Nilsen et al., 2009). However, despite a lower prevalence of disorders, little differences have been found in exposure-outcome associations in MoBa compared to the total Norwegian population (Nilsen et al., 2009).

Using the large, prospective population-based MoBa pregnancy cohort with longitudinal questionnaire follow-up and linkage to diagnoses from the nationwide patient registry, we identified specific mental traits (negative emotionality, emotional problems, behavioral problems) in childhood that were longitudinally associated with adolescent diagnosis of mood and anxiety disorders. The adolescence disorders were further associated with specific developmental profiles of combined emotional and behavioral problems. Our profile most predictive of mood or anxiety disorder was coherent with symptoms of disruptive mood dysregulation disorder, emphasizing its relevance to emotional disorders and the importance of a broad developmental perspective when evaluating risk of mood and anxiety disorders. Overall, our findings support a role of emotional dysregulation in the development of mood and anxiety disorders, highlighting the importance of childhood as a formative period, and suggesting potential for prevention and early intervention initiatives.

## Supporting information

STROBE checklist for cohort studies

## Data Availability

Data from the Norwegian Mother, Father and Child Cohort Study and the Medical Birth Registry of Norway used in this study are managed by the national health register holders in Norway (Norwegian Institute of public health) and can be made available to researchers, provided approval from the Regional Committees for Medical and Health Research Ethics (REC), compliance with the EU General Data Protection Regulation (GDPR) and approval from the data owners. The consent given by the participants does not open for storage of data on an individual level in repositories or journals. Researchers who want access to data sets for replication should apply through helsedata.no. Access to data sets requires approval from The Regional Committee for Medical and Health Research Ethics in Norway and an agreement with MoBa.

## Acknowledgements

We are grateful to all the participating families in Norway who take part in the Norwegian Mother, Father and Child Cohort Study (MoBa). MoBa is supported by the Norwegian Ministry of Health and Care Services and the Ministry of Education and Research. The current study was supported by grants from the Research Council of Norway (271555/F21, 273291, 223273, 274611, 288083), the Southern and Eastern Norway Regional Health Authority (2020022, 2018059, 2022083, 2019097,2018058) and facilities from the University of Oslo Faculty of Medicine. The work was partly performed on the TSD (Tjeneste for Sensitive Data), owned by the University of Oslo, operated, and developed by the TSD service group at the University of Oslo, IT-Department (USIT).

Key points

- Depression and anxiety account for a large global burden of disability in the adolescent and adult population. Still, there is limited knowledge of how and when early signs related to these disorders first manifest and develop.
- Our study indicates that childhood negative emotionality, behavioral and emotional difficulties are consistently and increasingly associated to adolescent anxiety and depression, with differences present already from 6 months of age.
- Our findings indicate that adolescent mood and anxiety disorders are often preceded by a gradual development of emotional and behavioral problems in childhood. Emotional dysregulation is highlighted as an important characteristic in the development of adolescent mood and anxiety disorders, with potential implications for the development and targeting of preventive interventions.

