## Supporting information for "Childhood temperamental, emotional, and behavioral predictors of clinical mood and anxiety disorders in adolescence"

### **Figure S1:** Flow diagram of included participants

No linkage to registry data in adolescence (age 10-18 years) (n=3,959)

Study population MoBa (n=114,326)

Study sample (n=110,367)

| Only dep (n=644) | Only anx (n=1,423) | Anx and dep (n=259) | Other (n=1,013) |
| --- | --- | --- | --- |

Developmental trajectories of mental health problems (n=81,886)

Emotional and behavioral problems (n=110,367)

Temperament and personality (n=110,367)

Any emotional disorder (n=3,339)

No emotional disorder (n=107,028)

8 yr (n=41,980)

5yr (n=39,645)

3yr (n=56,741)

18mnd (n=73,902)

6mnd (n=86,628)

**Figure S2** Correlation matrix for scales measuring mental health traits

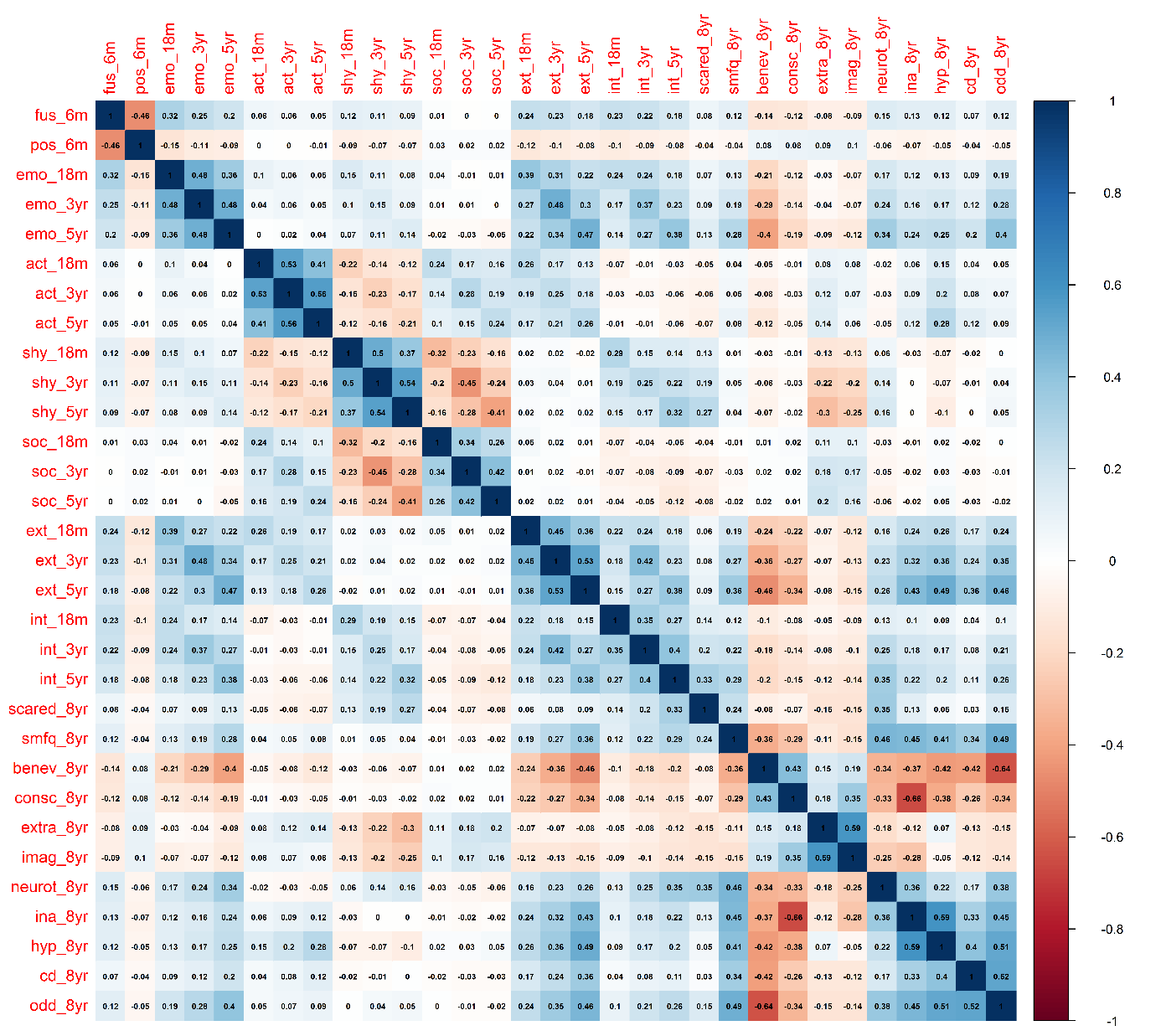

*Note: Pearson corelation coefficient for all childhood mental trait measures.*

**Figure S3** Strongest correlations for infant measure of fussy temperament, illustrating continuity for phenotype of negative emotionality.

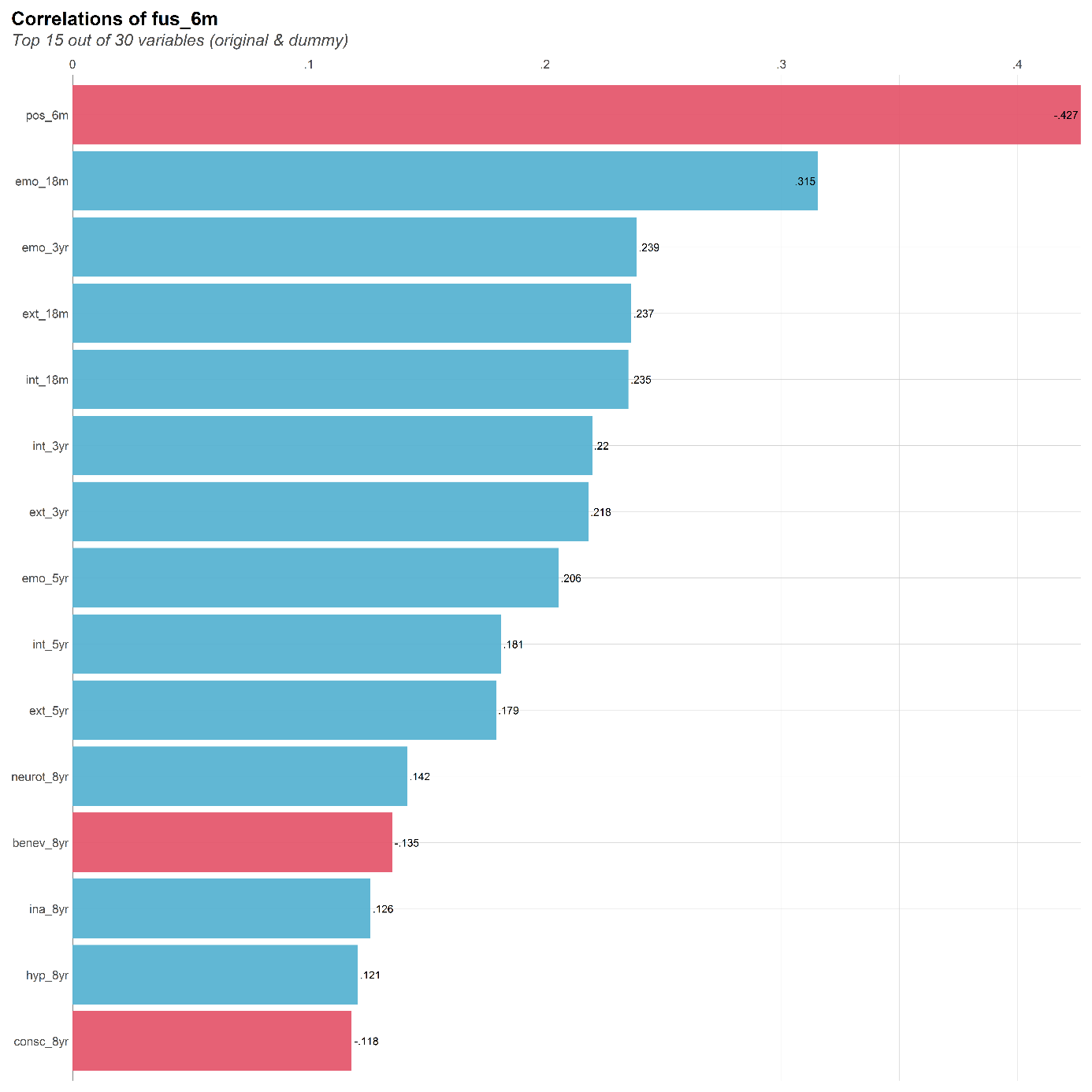

### **Table S1** Items included in each scale (childhood mental trait measure).

| **Instrument** | **Scale level data used in regression analyses** | **Item level data for each measure** |
| --- | --- | --- |
| **ICQ-6** | **Say whether you agree or disagree with the following statements about the child’s mood and temperament - how it is on a daily basis** | |
|  | Fussy temperament 6mths | - The child cries and complains a lot - The child is easy to calm when he/she cries (reversed) - The child is easily upset - When the baby cries, he/she usually cries loudly and vigorously - The child is so demanding that it would represent a considerable problem for most parents - The child requires a lot of attention - When left alone, he/she usually plays alone and is contented (reversed) |
|  | Positive temperament 6mths | - The child is easy to handle - The child smiles and laughs frequently |
| **EAS** | **To what extent do the following statements apply to your child's behavior during the last two month?** | |
|  | Emotionality 18mths | - Your child cries easily - Your child gets upset or sad easily - Your child reacts intensely when upset |
|  | Emotionality 3yrs | - Your child cries easily - Your child gets upset or sad easily - Your child reacts intensely when upset |
|  | Emotionality 5yrs | - Your child cries easily - Your child gets upset or sad easily - Your child reacts intensely when upset |
|  | Shyness 18mths | - Your child takes a long time to warm up to strangers - Your child is friendly towards and trusting of strangers (reversed) - Your child is very sociable (reversed) |
|  | Shyness 3yrs | - Your child takes a long time to warm up to strangers - Your child is friendly towards and trusting of strangers (reversed) - Your child is very sociable (reversed) |
|  | Shyness 5yrs | - Your child takes a long time to warm up to strangers - Your child is friendly towards and trusting of strangers (reversed) - Your child is very sociable (reversed) |
|  | Sociability 18mths | - Your child prefers playing with others rather than alone - Your child likes to be with people |
|  | Sociability 3yrs | - Your child prefers playing with others rather than alone - Your child finds other people more fun than anything else - Your child likes to be with people |
|  | Sociability 5yrs | - Your child prefers playing with others rather than alone - Your child finds other people more fun than anything else - Your child likes to be with people |
|  | Activity 18mths | - Your child is off and running as soon as he/she wakes up in the morning - Your child is always on the go - Your child prefers quiet, inactive games to more active ones(reversed) |
|  | Activity 3yrs | - Your child is off and running as soon as he/she wakes up in the morning - Your child is always on the go - Your child prefers quiet, inactive games to more active ones(reversed) |
|  | Activity 5yrs | - Your child is off and running as soon as he/she wakes up in the morning - Your child is always on the go - Your child prefers quiet, inactive games to more active ones(reversed) |
| **NHiPIC-30** | **Think back over the last year. How well do these statements apply to your child’s behavior over the past year?** | |
|  | Neuroticism 8yrs | - Is easily caught up in problems/Become easily panic - Is quick to worry about things - Doubt himself/herself - Is readily discouraged by imminent failure - Is quick to doubt his/her own capacities - Has confidence in own abilities (reversed)/ Feel at ease with him/herself(reversed) |
|  | Agreeableness 8yrs | - Obeys without protests - Takes himself/herself into consideration first(reversed) - Does everything to get his/her own way (reversed) - Imposes her or his will (reversed) - Is easily incensed by things (reversed) - Doesn’t envy others |
|  | Conscientiousness 8yrs | - Makes an all-out effort - Forgets anything and everything(reversed) - Prefers to leave work to others(reversed) - Is not very thorough - Finishes tasks to the very end - Carries out work to the last detail |
|  | Imagination 8yrs | - Has a broad range of interests - Derives pleasure from creating things - Is quick to understands things - Has a rich imagination - Is interested in all that is new (is interested in anything) - Can express himself/herself well |
|  | Extraversion 8yrs | - Is chatty - Enjoys life - Has an infectious laugh - Talks about own feelings - Is constantly on the move - Talks to people easily |
| **CBCL** | **To what extent are the following statements true of your child’s behavior during the last two months?** | |
|  | Emotional problems 18 mths | - Disturbed by any change in routine - Clings to adults or too dependent - Gets too upset when separated from parents - Too fearful or anxious - Doesn’t eat well |
|  | Emotional problems 3yrs | - Disturbed by any change in routine - Sudden changes in moods or feelings - Clings to adults or too dependent - Gets too upset when separated from parents - Too fearful or anxious - Constipated, doesn’t move bowels - Doesn’t eat well - Stomach aches or cramps (without medical cause) - Vomiting, throwing up (without medical cause) |
|  | Emotional problems 5 yrs | - Disturbed by any change in routine - Clings to adults or too dependent - Gets too upset when separated from parents - Too fearful or anxious - Nervous, highstrung, or tense - Self-conscious or easily embarrassed - Unhappy, sad or depressed - Feelings are easily hurt - Doesn’t eat well - Stomach aches or cramps (without medical cause) - Vomiting, throwing up (without medical cause) |
|  | Behavioral problems 18 mths | - Can’t concentrate, can’t pay attention for long - Can’t sit still, restless or overactive - Quickly shifts from one activity to another - Hits others - Defiant - Doesn’t seem to feel guilty after misbehaving - Punishment doesn’t change his/her behavior - Gets in many fights |
|  | Behavioral problems 3yrs | - Can’t concentrate, can’t pay attention for long - Can’t sit still, restless or overactive - Poorly coordinated or clumsy - Quickly shifts from one activity to another - Defiant - Can’t stand waiting, wants everything now - Demands must be met immediately - Doesn’t seem to feel guilty after misbehaving - Gets in many fights - Hits others - Punishment doesn’t change his/her behavior |
|  | Behavioral problems 5 yrs | - Poorly coordinated or clumsy - Quickly shifts from one activity to another - Can’t sit still, restless or overactive - Quickly shifts from one activity to another - Can’t stand waiting, wants everything now - Defiant - Demands must be met immediately - Doesn’t seem to feel guilty after misbehaving - Gets in many fights - Hits others |
| **SMFQ** | **Mark how true each item has been for your child during the two last weeks.** | |
|  | Depressive symptoms 8yrs | - Felt miserable or unhappy - Felt so tired that s/he just sat around and did nothing - Was very restless - Didn’t enjoy anything at all - Felt s/he was no good anymore - Cried a lot - Hated him/herself - Thought s/he could never be as good as other kids - Felt lonely - Thought nobody really loved him/her - Felt s/he was a bad person - Felt s/he did everything wrong - Found it hard to think/concentrate |
| **SCARED** | **The questions below are about how your child have felt or behaved recently** | |
|  | Anxiety symptoms 8yrs | - My child gets really frightened for no reason at all - My child is afraid to be alone in the house - People tell my child that he/she worries too much - My child is scared to go to school - My child is shy |
| **RS-DBD** | **Mark the box that best describes your child’s behavior during the last 12 months/last year** | |
|  | Hyperactivity 8yrs | - Fidgets with hands or feet or squirms in seat (sits uneasily) - Leaves seat in classroom or in other situations in which remaining seated is expected (e.g. at the table or in group gathering) - Runs about or climbs excessively in situations in which it is inappropriate - Has difficulty playing or engaging in leisure activities quietly - Is “on the go” or acts as if “driven by a motor” - Talks excessively - Blurts out answers before questions have been completed - Has difficulty awaiting turn - Interrupts or intrudes on others, such as in conversation or play |
|  | Inattention 8yrs | - Fails to give close attention to details or makes careless mistakes in schoolwork - Has difficulty sustaining attention in tasks or play activities - Does not seem to listen when spoken to directly - Does not follow through on instructions and fails to finish school work, chores or duties (not due to oppositional behaviour or failure to understand instructions) - Has difficulty organizing tasks and activities - Avoids, dislikes or is reluctant to engage in tasks that require sustained mental effort (such as schoolwork or homework) - Loses things necessary for tasks or activities (pencils, books, toys) - Is easily distracted - Is forgetful in daily activities |
|  | Oppositional defiant disorder symptoms 8yrs | - Loses temper (tantrums) - Argues with adults - Actively defies or refuses to comply with adults’ requests or rules - Deliberately annoys people - Blames others for his/her mistakes or misbehaviour - Is touchy or easily annoyed by others - Is angry and resentful - Is spiteful or vindictive |
|  | Conduct disorder symptoms 8yrs | - Bullies, threatens or intimidates others - Initiates physical fights - Has been physically cruel to others - Has harassed or injured animals physically - Has stolen items of nontrivial value without confronting a victim (e.g. shoplifting) - Has deliberately destroyed other’s property - Has been truant from school - Has used an object that can cause serious physical harm to others (e.g. a bat, stone, knife, heavy toy) |
| **CBCL repeated measures included in growth models** | **To what extent are the following statements true of your child’s behavior during the last two months?** | |
|  | Emotional problems (repeated measures) | - Disturbed by any change in routine - Clings to adults or too dependent - Gets too upset when separated from parents - Too fearful or anxious - Doesn’t eat well |
|  | Behavioral problems (repeated measures) | - Can’t concentrate, can’t pay attention for long - Cant sit still, restless or overactive - Hits others - Doesn’t seem to feel guilty after misbehaving - Gets in many fights - Quickly shifts from one activity to another - Defiant - Punishment doesn’t change his/her behaviour |

### **Table S2** Ordinal Cronbach’s alpha for scales measuring mental health traits used in analyses

| **Key study variables** | **Ordinal Cronbach alpha** |
| --- | --- |
| Fussy temperament 6mths | 0.81 |
| Positive temperament 6mths | 0.70 |
| Emotionality 18mths | 0.69 |
| Emotionality 3yrs | 0.69 |
| Emotionality 5yrs | 0.80 |
| Shyness 18mths | 0.70 |
| Shyness 3yrs | 0.71 |
| Shyness 5yrs | 0.76 |
| Sociability 18mths | 0.43 |
| Sociability 3yrs | 0.61 |
| Sociability 5yrs | 0.81 |
| Activity 18mths | 0.72 |
| Activity 3yrs | 0.69 |
| Activity 5yrs | 0.75 |
| Neuroticism 8yrs | 0.83 |
| Agreeableness 8yrs | 0.81 |
| Conscientiousness 8yrs | 0.79 |
| Imagination 8yrs | 0.74 |
| Extraversion 8yrs | 0.70 |
| Emotional problems 18 mths | 0.66 |
| Emotional problems 3yrs | 0.74 |
| Emotional problems 5 yrs | 0.85 |
| Behavioral problems 18 mths | 0.69 |
| Behavioral problems 3yrs | 0.80 |
| Behavioral problems 5 yrs | 0.83 |
| Depressive symptoms 8yrs | 0.92 |
| Anxiety symptoms 8yrs | 0.76 |
| Hyperactivity 8yrs | 0.91 |
| Inattention 8yrs | 0.92 |
| Oppositional defiant disorder symptoms 8yrs | 0.91 |
| Conduct disorder symptoms 8yrs | 0.88 |
| Emotional problems development 18 mths | 0.66 |
| Emotional problems development 3yrs | 0.68 |
| Emotional problems development 5 yrs | 0.73 |
| Behavioral problems development 18 mths | 0.69 |
| Behavioral problems development 3yrs | 0.77 |
| Behavioral problems development 5 yrs | 0.81 |

### **Supplementary text1**

**Analytical tools used in analyses:** R-version 4.0.3 was used for all analysis except parts of the developmental models utilizing M-plus version 8.3(Muthén LK & Muthén B.O, 1998-2017). The R-package “phenotools” (https://github.com/psychgen/phenotools), developed for analysis of MoBa-questionnaire data and linked registry data, was used to calculate scores for the scales included and prepare diagnostic data. In this tool scale scores used in presented analyses were computed as the mean of all included items, multiplied by number of items included in the scale. Scores were calculated only for individuals with values registered on more than 50% of the scale items (mother-reported), else NA was set for the individual.

Descriptive analyses were conducted with the R-package “psych” version 2.1.3 (Revelle W, 2021). Logistic regression was conducted using “miceadds” version 3.11-6 (Robitzsch A & Grund S, 2021) and “gtsummary” version 1.4.2 (Sjoberg D, Whiting K, Curry M, Lavery J& Larmarange J, 2021) combined with “jtools” version 2.1.1(Long JA, 2020). The latent profile analysis (LPA) was conducted via the R-package “MplusAutomation” version 1.0.0 (Hallquist MN & Wiley JF, 2017), depending on Mplus software version 8.3.

### **Figure S4:** Latent profile analysis incorporating latent growth models for developmental profiles of emotional and behavioral problems

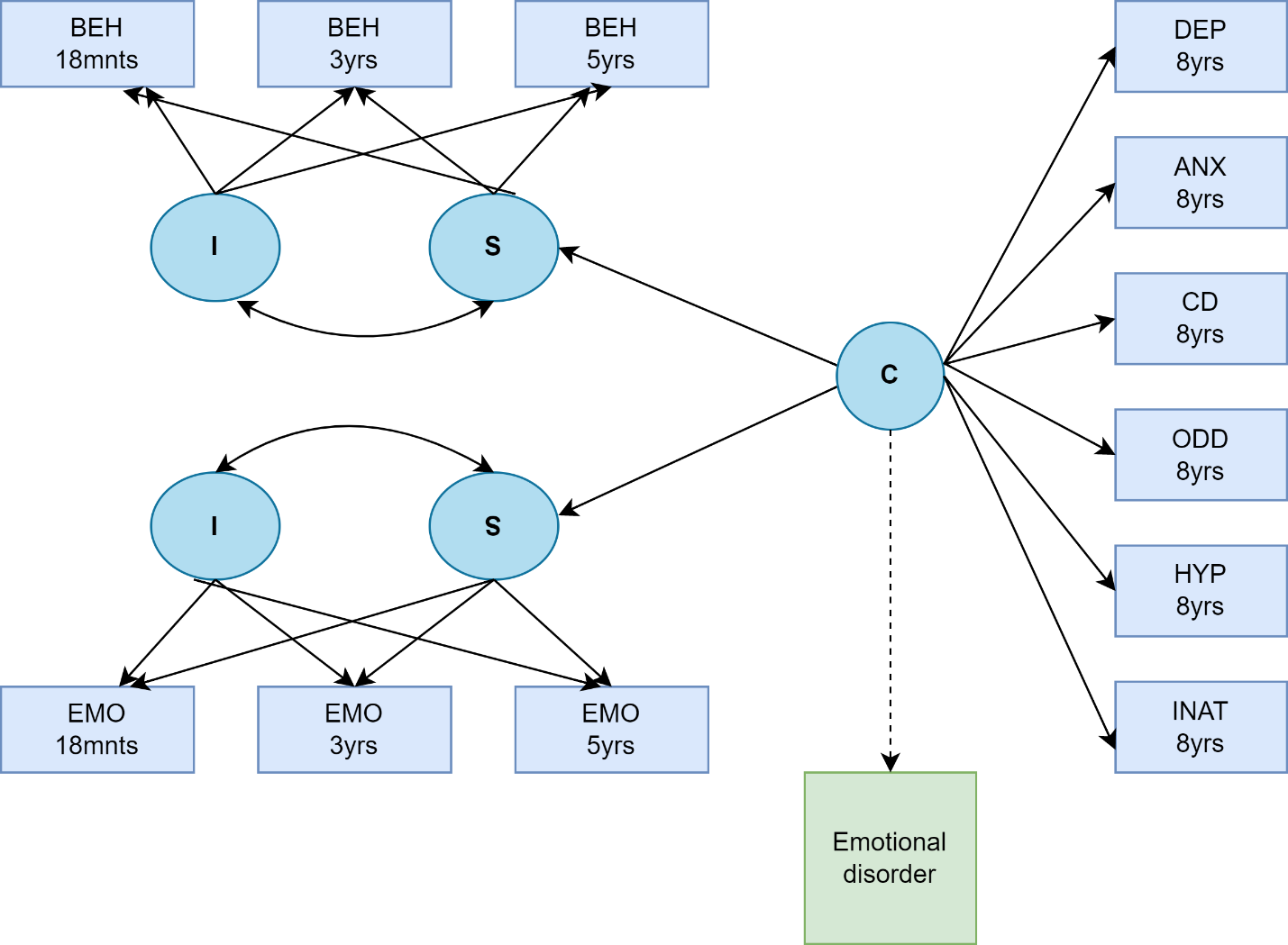

**Note**: Illustration of methods for developmental profiles incorporating latent growth models in 3-step maximum likelihood latent profile analysis. Boxes represent observed variables and circles model-estimated latent variables; I = intercept factor, loads equally on observed variables at all waves; C = categorical latent variable, subdividing the sample into a specified number of classes according to values: S = slope factor with loadings 0(18mnts), 1.5(3yrs), and 3.5(5yrs), corresponding to temporal distance between the measurements; BEH = Behavioral problems; EMO = Emotional problems; DEP= Depressive symptoms; ANX = Anxiety symptoms; CD= Conduct disorder symptoms; ODD = Oppositional defiant disorder symptoms; HYP = Hyperactivity; INAT= Inattention. Emotional/ behavioral intercept/slope variables and 8-year observed variables are intercorrelated within class (paths omitted from diagram for clarity) in step 1 of the latent profile analysis. Emotional disorder is included as distal outcome in step 3 in model-specification (indicated by dashed line).

For further methodological details on latent growth models see “Genetic liability for schizophrenia and childhood psychopathology in the general population” by L.J. Hannigan in Schizophrenia Bulletin Volume 47, Issue 4, July 2021, Pages 1179–1189 (Hannigan LJ, et al., 2021).

### **Table S3**: Comparing attrition between diagnosed and non-diagnosed individuals.

|  | **Emotional disorder**  **(n=3,339)** | **No emotional disorder (n=107,028)** | **Total (n=110,367)** |
| --- | --- | --- | --- |
| **Demographic information** | **n Missing, (% total)** | **n Missing, (% total)** | **n Missing, (% total)** |
| Sex, male | 0(0%) | <5 (<0.1%) | <5 (<0.1%) |
| Age end of follow-up in 2018, years | 0(0%) | 0(0%) | 0(0%) |
| Age first emotional disorder, years | 0(0%) | 0(0%) | 0(0%) |
| Parental education level | 595 (18%) | 15,843 (15%) | 16,438 (15%) |
| Marital status | 0(0%) | <5 (<0.1%) | <5 (<0.1%) |
| Maternal age | <5 (<0.1%) | 52 (<0.1%) | 55 (<0.1%) |
| Parity | 0(0%) | <5 (<0.1%) | <5 (<0.1%) |
| Mother’s country of birth | 65(1.9%) | 1,907 (1.8%) | 1,972 (1.8%) |
| **Co-occurring diagnoses** |  |  |  |
| ADHD, n, (%) | 0(0%) | 0(0%) | 0(0%) |
| OCD, n, (%) | 0(0%) | 0(0%) | 0(0%) |
| CD, n, (%) | 0(0%) | 0(0%) | 0(0%) |
| **Key study variables** |  |  |  |
| Fussy temperament 6mths | 741 (22%) | 23,424 (22%) | 24,165 (22%) |
| Positive temperament 6mths | 742 (22%) | 23,375 (22%) | 24,117 (22%) |
| Emotionality 18mths | 1,138 (34%) | 35,936 (34%) | 37,074 (34%) |
| Emotionality 3yrs | 1,776 (53%) | 52,144 (49%) | 53,920 (49%) |
| Emotionality 5yrs | 2,564 (77%) | 68,376 (64%) | 70,940 (64%) |
| Shyness 18mths | 1,137 (34%) | 35,900 (34%) | 37,037 (34%) |
| Shyness 3yrs | 1,777 (53%) | 52,132 (49%) | 53,909 (49%) |
| Shyness 5yrs | 2,566 (77%) | 68,406 (64%) | 70,972 (64%) |
| Sociability 18mths | 1,157 (35%) | 36,393 (34%) | 37,550 (34%) |
| Sociability 3yrs | 1,775 (53%) | 52,123 (49%) | 53,898 (49%) |
| Sociability 5yrs | 2,563 (77%) | 68,368 (64%) | 70,931 (64%) |
| Activity 18mths | 1,137 (34%) | 35,860 (34%) | 36,997 (34%) |
| Activity 3yrs | 1,775 (53%) | 52,103 (49%) | 53,878 (49%) |
| Activity 5yrs | 2,563 (77%) | 68,366 (64%) | 70,929 (64%) |
| Neuroticism 8yrs | 2,320 (69%) | 66,247 (62%) | 68,567 (62%) |
| Agreeableness 8yrs | 2,318 (69%) | 66,226 (62%) | 68,544 (62%) |
| Conscientiousness 8yrs | 2,317 (69%) | 66,223 (62%) | 68,540 (62%) |
| Imagination 8yrs | 2,320 (69%) | 66,234 (62%) | 68,554 (62%) |
| Extraversion 8yrs | 2,320 (69%) | 66,239 (62%) | 68,559 (62%) |
| Emotional problems 18 mths | 1,478 (44%) | 40,302 (38%) | 41,780 (38%) |
| Emotional problems 3yrs | 1,778 (53%) | 52,257 (49%) | 54,035 (49%) |
| Emotional problems 5 yrs | 2,562 (77%) | 68,423 (64%) | 70,985 (64%) |
| Behavioral problems 18 mths | 1,125 (34%) | 35,691 (33%) | 36,816 (33%) |
| Behavioral problems 3yrs | 1,778 (53%) | 52,259 (49%) | 54,037 (49%) |
| Behavioral problems 5 yrs | 2,561 (77%) | 68,414 (64%) | 70,975 (64%) |
| Depressive symptoms 8yrs | 2,322 (70%) | 66,246 (62%) | 68,568 (62%) |
| Anxiety symptoms 8yrs | 2,321 (70%) | 66,165 (62%) | 68,486 (62%) |
| Hyperactivity 8yrs | 2,320 (69%) | 66,206 (62%) | 68,526 (62%) |
| Inattention 8yrs | 2,320 (69%) | 66,198 (62%) | 68,518 (62%) |
| Oppositional defiant disorder symptoms 8yrs | 2,320 (69%) | 66,210 (62%) | 68,530 (62%) |
| Conduct disorder symptoms 8yrs | 2,321 (70%) | 66,163 (62%) | 68,484 (62%) |

**Note:** Co-occurring diagnoses= additional diagnosis during follow-up. ADHD= Attention Deficit Hyperactivity Disorder. OCD= obsessive-compulsive disorder. CD= Conduct disorders.

### **Table S4**: Evaluating demographic characteristics and descriptive information for key study variables for individuals included and not included in trajectory models

|  | **Individuals included in developmental models (n=81,886)** | **Individuals with insufficient data for developmental models (n=28,480)** | **Total sample**  **(n=110,367)** |
| --- | --- | --- | --- |
| Sex, male, n (%) | 41931 (51.2%) | 14579 (51.2%) | 56510 (51.2%) |
| Age end of follow-up, years, *m* (SD) | 13.05 (2.11) | 13.01 (2.19) | 13.04 (2.13) |
| Age first emotional disorder, years, M, (SD) | 11.31 (3.13) | 11.28 (3.31) | 11.30 (3.18) |
| Highly educated mother or father, n (%) | 57021 (74.7%) | 11069 (62.9%) | 68090 (72.5%) |
| Marital status, n (%)^a^ | 78949 (96.4%) | 26508 (93.1%) | 4909 (4.4%) |
| Maternal age, m, (SD)^b^ | 30.29 (4.50) | 29.65 (4.99) | 30.12 (4.64) |
| Parity, n (%) primiparous | 37298 (45.5%) | 11041 (38.8%) | 48339 (43.8%) |
| **Mother’s country of birth** |  |  |  |
| Norway, n(%) | 74788 (93.0%) | 24794 (88.6%) | 99582 (91.9%) |
| Other high-income-country, n (%) | 3623 (4.5%) | 1314 (4.7%) | 4937 (4.6%) |
| Other GDB 7 super region country, n(%) | 2011 (2.5%) | 1865 (6.7%) | 3876 (3.6%) |
| **Diagnostic status** |  |  |  |
| Emotional disorder, n(%) | 2421 (3.0%) | 918 (3.2%) | 3339 (3.0%) |
| ADHD, n, (%) | 3156 (3.9%) | 1393 (4.9%) | 4549 (4.1%) |
| OCD, n, (%) | 335 (0.4%) | 126 (0.4%) | 461 (0.4%) |
| CD, n, (%) | 425 (0.5%) | 163 (0.6%) | 588 (0.5%) |
| **Key study variables, M, (SD)** |  |  |  |
| Fussy temperament 6mths | 9.40 (5.61) | 9.57 (5.75) | 9.42 (5.62) |
| Positive temperament 6mths | 11.06 (1.62) | 11.04 (1.69) | 11.06 (1.63) |
| Emotionality 18mths | 5.22 (2.30) | 5.30 (2.31) | 5.22 (2.30) |
| Emotionality 3yrs | 5.38 (2.31) | 6.33 (2.55) | 5.38 (2.31) |
| Emotionality 5yrs | 4.25 (2.50) | 3.60 (0.89) | 4.25 (2.50) |
| Shyness 18mths | 3.14 (1.93) | 3.10 (1.98) | 3.14 (1.93) |
| Shyness 3yrs | 3.65 (2.04) | 3.88 (2.31) | 3.65 (2.04) |
| Shyness 5yrs | 3.26 (2.14) | 3.20 (3.56) | 3.26 (2.14) |
| Sociability 18mths | 8.90 (1.53) | 8.75 (1.92) | 8.90 (1.53) |
| Sociability 3yrs | 8.04 (1.71) | 8.12 (1.88) | 8.04 (1.71) |
| Sociability 5yrs | 9.21 (1.91) | 9.17 (2.23) | 9.21 (1.91) |
| Activity 18mths | 9.06 (1.96) | 8.36 (2.26) | 9.06 (1.96) |
| Activity 3yrs | 7.87 (2.11) | 7.73 (2.65) | 7.87 (2.11) |
| Activity 5yrs | 6.68 (2.15) | 5.67 (0.52) | 6.68 (2.15) |
| Neuroticism 8yrs | 7.58 (4.57) | NA | 7.58 (4.57) |
| Agreeableness 8yrs | 15.51 (3.75) | NA | 15.51 (3.75) |
| Conscientiousness 8yrs | 15.92 (3.64) | NA | 15.92 (3.64) |
| Imagination 8yrs | 17.85 (3.44) | NA | 17.85 (3.44) |
| Extraversion 8yrs | 16.38 (3.79) | NA | 16.38 (3.79) |
| Emotional problems 18 mths | 1.33 (1.24) | NA | 1.33 (1.24) |
| Emotional problems 3yrs | 2.22 (1.99) | NA | 2.22 (1.99) |
| Emotional problems 5 yrs | 2.03 (2.22) | NA | 2.03 (2.22) |
| Behavioral problems 18 mths | 3.95 (2.27) | NA | 3.95 (2.27) |
| Behavioral problems 3yrs | 5.54 (3.19) | NA | 5.54 (3.19) |
| Behavioral problems 5 yrs | 3.76 (3.07) | NA | 3.76 (3.07) |
| Depressive symptoms 8yrs | 1.87 (2.45) | NA | 1.87 (2.45) |
| Anxiety symptoms 8yrs | 1.03 (1.20) | NA | 1.03 (1.20) |
| Hyperactivity 8yrs | 3.56 (3.90) | NA | 3.56 (3.90) |
| Inattention 8yrs | 4.99 (4.15) | NA | 4.99 (4.15) |
| Oppositional defiant disorder symptoms 8yrs | 3.42 (3.16) | NA | 3.42 (3.16) |
| Conduct disorder symptoms 8yrs | 0.78 (1.51) | NA | 0.78 (1.51) |

**Note:**.. **a**:.n(%) married/registered partner/co-habitant. **b:** mean maternal age at pregnancy is calculated for mothers between age 17 and 45 due to lack of information on age for mothers above or below this age-range (N=58)

Co-occurring diagnoses= additional diagnosis during follow-up. ADHD= Attention Deficit Hyperactivity Disorder. OCD= obsessive-compulsive disorder. CD= Conduct disorders. Co-occurring diagnoses= additional diagnosis during follow-up.

### **Table S5** Descriptive statistics for main study variables

| **Key study variables, M, (SD)** | **Emotional disorder (n=3,339)** | **No emotional disorder (n=107,028)** | **Total (n=110,367)** |
| --- | --- | --- | --- |
| Fussy temperament 6mths | 9.79 (5.78) | 9.41 (5.62) | 9.42 (5.62) |
| Positive temperament 6mths | 11.06 (1.59) | 11.06 (1.63) | 11.06 (1.63) |
| Emotionality 18mths | 5.40 (2.33) | 5.22 (2.30) | 5.22 (2.30) |
| Emotionality 3yrs | 5.79 (2.39) | 5.37 (2.31) | 5.38 (2.31) |
| Emotionality 5yrs | 5.01 (2.68) | 4.24 (2.49) | 4.25 (2.50) |
| Shyness 18mths | 3.12 (1.99) | 3.14 (1.93) | 3.14 (1.93) |
| Shyness 3yrs | 3.70 (2.10) | 3.65 (2.04) | 3.65 (2.04) |
| Shyness 5yrs | 3.51 (2.36) | 3.26 (2.14) | 3.26 (2.14) |
| Sociability 18mths | 8.87 (1.61) | 8.90 (1.53) | 8.90 (1.53) |
| Sociability 3yrs | 7.96 (1.76) | 8.04 (1.71) | 8.04 (1.71) |
| Sociability 5yrs | 9.08 (2.03) | 9.22 (1.91) | 9.21 (1.91) |
| Activity 18mths | 9.17 (2.01) | 9.06 (1.96) | 9.06 (1.96) |
| Activity 3yrs | 7.98 (2.21) | 7.87 (2.10) | 7.87 (2.11) |
| Activity 5yrs | 6.86 (2.38) | 6.68 (2.14) | 6.68 (2.15) |
| Neuroticism 8yrs | 10.20 (5.30) | 7.51 (4.53) | 7.58 (4.57) |
| Conscientiousness 8yrs | 15.08 (4.03) | 15.95 (3.63) | 15.92 (3.64) |
| Agreeableness 8yrs | 14.16 (4.30) | 15.55 (3.73) | 15.51 (3.75) |
| Imagination 8yrs | 16.98 (3.89) | 17.87 (3.43) | 17.85 (3.44) |
| Extraversion 8yrs | 15.71 (4.03) | 16.40 (3.78) | 16.38 (3.79) |
| Emotional problems 18 mths | 1.52 (1.35) | 1.32 (1.24) | 1.33 (1.24) |
| Emotional problems 3yrs | 2.68 (2.19) | 2.21 (1.98) | 2.22 (1.99) |
| Emotional problems 5 yrs | 3.14 (3.08) | 2.01 (2.19) | 2.03 (2.22) |
| Behavioral problems 18 mths | 4.23 (2.40) | 3.94 (2.27) | 3.95 (2.27) |
| Behavioral problems 3yrs | 6.39 (3.54) | 5.51 (3.18) | 5.54 (3.19) |
| Behavioral problems 5 yrs | 5.08 (3.84) | 3.73 (3.05) | 3.76 (3.07) |
| Depressive symptoms 8yrs | 3.47 (3.77) | 1.83 (2.40) | 1.87 (2.45) |
| Anxiety symptoms 8yrs | 1.81 (1.84) | 1.01 (1.17) | 1.03 (1.20) |
| Inattention 8yrs | 6.62 (5.27) | 4.94 (4.11) | 4.99 (4.15) |
| Hyperactivity 8yrs | 5.07 (5.35) | 3.52 (3.85) | 3.56 (3.90) |
| Conduct disorder symptoms 8yrs | 1.32 (2.14) | 0.77 (1.49) | 0.78 (1.51) |
| Oppositional defiant disorder symptoms 8yrs | 5.08 (4.31) | 3.38 (3.12) | 3.42 (3.16) |

### **Table S6** Association between mental health traits in childhood and emotional disorder in adolescence (10-18 years).

| **Emotional disorder in adolescence (10-18 years)** | | | | |
| --- | --- | --- | --- | --- |
| **Temperament and personality traits** | **OR^1^** | **95% CI^1^** | **p-value** | **q-value^2^** |
| Fussy temperament 6mths | 1.07 | 1.03, 1.11 | <0.001 | <0.001 |
| Positive temperament 6mths | 1.00 | 0.96, 1.03 | 0.8 | 0.9 |
| Emotionality 18mths | 1.08 | 1.04, 1.13 | <0.001 | <0.001 |
| Emotionality 3yrs | 1.19 | 1.13, 1.25 | <0.001 | <0.001 |
| Emotionality 5yrs | 1.38 | 1.28, 1.48 | <0.001 | <0.001 |
| Shyness 18mths | 1.00 | 0.96, 1.04 | >0.9 | >0.9 |
| Shyness 3yrs | 1.04 | 0.99, 1.10 | 0.10 | 0.11 |
| Shyness 5yrs | 1.14 | 1.06, 1.23 | <0.001 | 0.001 |
| Sociability 18mths | 0.97 | 0.93, 1.01 | 0.2 | 0.2 |
| Sociability 3yrs | 0.95 | 0.90, 1.00 | 0.067 | 0.082 |
| Sociability 5yrs | 0.94 | 0.87, 1.01 | 0.083 | 0.10 |
| Activity 18mths | 1.06 | 1.02, 1.11 | 0.008 | 0.011 |
| Activity 3yrs | 1.06 | 1.00, 1.11 | 0.042 | 0.054 |
| Activity 5yrs | 1.08 | 1.00, 1.17 | 0.046 | 0.058 |
| Neuroticism 8yrs | 1.81 | 1.71, 1.92 | <0.001 | <0.001 |
| Conscientiousness 8yrs | 0.79 | 0.74, 0.84 | <0.001 | <0.001 |
| Agreeableness 8yrs | 0.71 | 0.66, 0.75 | <0.001 | <0.001 |
| Imagination 8yrs | 0.80 | 0.75, 0.86 | <0.001 | <0.001 |
| Extraversion 8yrs | 0.84 | 0.79, 0.90 | <0.001 | <0.001 |
| **Mental health problems** |  |  |  |  |
| Emotional problems 18 mths | 1.16 | 1.11, 1.21 | <0.001 | <0.001 |
| Emotional problems 3yrs | 1.22 | 1.17, 1.28 | <0.001 | <0.001 |
| Emotional problems 5 yrs | 1.50 | 1.43, 1.57 | <0.001 | <0.001 |
| Behavioral problems 18 mths | 1.12 | 1.07, 1.16 | <0.001 | <0.001 |
| Behavioral problems 3yrs | 1.28 | 1.22, 1.35 | <0.001 | <0.001 |
| Behavioral problems 5 yrs | 1.45 | 1.36, 1.54 | <0.001 | <0.001 |
| Depressive symptoms 8yrs | 1.51 | 1.46, 1.58 | <0.001 | <0.001 |
| Anxiety symptoms 8yrs | 1.62 | 1.55, 1.70 | <0.001 | <0.001 |
| Inattention 8yrs | 1.39 | 1.32, 1.45 | <0.001 | <0.001 |
| Hyperactivity 8yrs | 1.37 | 1.30, 1.44 | <0.001 | <0.001 |
| Conduct disorder symptoms 8yrs | 1.30 | 1.24, 1.36 | <0.001 | <0.001 |
| Oppositional defiant disorder symptoms 8yrs | 1.51 | 1.44, 1.58 | <0.001 | <0.001 |
| ^1^OR = Odds Ratio per standard deviation change in trait score, CI = Confidence Interval | | | | |
| ^2^False discovery rate correction for multiple testing. Q-value calculated for all 31 evaluated measures (temperament traits and emotional symptoms). | | | | |
| OR for each scale is calculated separately in logistic regression with birth year and sex as covariates. Calculated using robust standard errors HC0 and clustered for maternal ID. | | | | |

### **Table S7** Analysis illustrating difference in odds ratio for anxiety and depression in adolescence (10-18 years) per standard deviation change in childhood mental health traits.

|  | **Depressive disorder** | | **Anxiety disorder** | |
| --- | --- | --- | --- | --- |
| **Characteristic** | **OR^1^** | **95% CI^1^** | **OR^1^** | **95% CI^1^** |
| Fussy temperament 6mths | 1.02 | 0.93, 1.13 | 1.06 | 1.00, 1.12 |
| Positive temperament 6mths | 0.99 | 0.90, 1.08 | 1.01 | 0.95, 1.07 |
| Emotionality 18mths | 1.06 | 0.97, 1.17 | 1.07 | 1.01, 1.14 |
| Emotionality 3yrs | 1.12 | 0.99, 1.26 | 1.21 | 1.12, 1.31 |
| Emotionality 5yrs | 1.13 | 0.90, 1.41 | 1.31 | 1.18, 1.46 |
| Shyness 18mths | 0.98 | 0.89, 1.08 | 1.03 | 0.96, 1.10 |
| Shyness 3yrs | 0.92 | 0.82, 1.04 | 1.13 | 1.05, 1.22 |
| Shyness 5yrs | 1.02 | 0.81, 1.29 | 1.19 | 1.07, 1.33 |
| Sociability 18mths | 0.95 | 0.86, 1.04 | 1.00 | 0.93, 1.07 |
| Sociability 3yrs | 0.89 | 0.78, 1.02 | 0.98 | 0.90, 1.06 |
| Sociability 5yrs | 0.84 | 0.68, 1.03 | 1.03 | 0.93, 1.15 |
| Activity 18mths | 1.04 | 0.94, 1.15 | 1.04 | 0.97, 1.11 |
| Activity 3yrs | 1.00 | 0.87, 1.13 | 1.01 | 0.94, 1.10 |
| Activity 5yrs | 0.85 | 0.68, 1.07 | 1.11 | 0.99, 1.24 |
| Neuroticism 8yrs | 1.33 | 1.14, 1.55 | 1.87 | 1.71, 2.04 |
| Conscientiousness 8yrs | 0.85 | 0.72, 1.01 | 0.80 | 0.73, 0.88 |
| Agreeableness 8yrs | 0.84 | 0.71, 0.99 | 0.78 | 0.71, 0.85 |
| Imagination 8yrs | 0.93 | 0.78, 1.11 | 0.75 | 0.68, 0.81 |
| Extraversion 8yrs | 0.82 | 0.69, 0.98 | 0.85 | 0.77, 0.93 |
| Emotional problems 18 mths | 0.99 | 0.89, 1.11 | 1.19 | 1.12, 1.27 |
| Emotional problems 3yrs | 1.05 | 0.92, 1.19 | 1.24 | 1.16, 1.31 |
| Emotional problems 5 yrs | 1.35 | 1.14, 1.59 | 1.51 | 1.42, 1.62 |
| Behavioral problems 18 mths | 1.01 | 0.92, 1.11 | 1.10 | 1.03, 1.17 |
| Behavioral problems 3yrs | 1.12 | 0.99, 1.27 | 1.22 | 1.13, 1.31 |
| Behavioral problems 5 yrs | 1.36 | 1.11, 1.65 | 1.29 | 1.17, 1.41 |
| Depressive symptoms 8yrs | 1.39 | 1.27, 1.51 | 1.40 | 1.33, 1.49 |
| Anxiety symptoms 8yrs | 1.00 | 0.84, 1.19 | 1.80 | 1.70, 1.91 |
| Inattention 8yrs | 1.17 | 1.03, 1.34 | 1.32 | 1.23, 1.42 |
| Hyperactivity 8yrs | 1.14 | 0.98, 1.32 | 1.27 | 1.18, 1.37 |
| Conduct disorder symptoms 8yrs | 1.17 | 1.06, 1.31 | 1.14 | 1.06, 1.23 |
| Oppositional defiant disorder symptoms 8yrs | 1.35 | 1.19, 1.52 | 1.33 | 1.23, 1.43 |
| ^1^OR = Odds Ratio per standard deviation change in trait score, CI = Confidence Interval | | | | |
| OR for each scale is calculated separately in logistic regression with birth year and sex as covariates. Q-value calculated for all 31 evaluated measures (temperament traits and emotional symptoms). Calculated using robust standard errors HC0 and clustered for maternal ID. | | | | |

### **Table S8:** Fit statistics for evaluated developmental models

| **Number of profiles in model** | **LL** | **BIC** | **aBIC** | **AIC** | **AICc** | **Entropy** | **VLMR** | **VLMR_P-value** |
| --- | --- | --- | --- | --- | --- | --- | --- | --- |
| 1 | -1186738 | 2374009 | 2373860 | 2373571 | 2373571 | NA | NA | NA |
| 2 | -1173971 | 2348598 | 2348414 | 2348058 | 2348058 | 0.808 | 25535 | 0 |
| 3 | -1167206 | 2335192 | 2334973 | 2334550 | 2334550 | 0.783 | 13530 | 0 |
| 4 | -1160850 | 2322605 | 2322350 | 2321860 | 2321860 | 0.756 | 11966 | 0 |
| 5 | -1156767 | 2314565 | 2314276 | 2313717 | 2313717 | 0.754 | 8164 | 0 |
| 6 | -1152471 | 2306096 | 2305771 | 2305146 | 2305146 | 0.753 | 6670 | 0 |
| 7 | -1150175 | 2301629 | 2301270 | 2300577 | 2300577 | 0.738 | 4590 | 0 |
| 8 | -1147816 | 2297036 | 2296642 | 2295881 | 2295882 | 0.756 | 4717 | 0.0831 |

**Note:** LL= Loglikelihood value for final model. BIC= Bayesian information criterion. aBIC=Sample Size Adjusted Bayesian information criterion. AIC= First-order Akaike Information Criterion. AICc= Second-order Akaike Information Criterion. VLMR= Vuong–Lo–Mendell–Rubin likelihood ratio test.

### **Table S9:** Demographic characteristics of each developmental profile

| **Characteristic** | **Profile 1**  **(n=3,087)** | **Profile 2**  **(n=69,522)** | **Profile 3**  **(n=4,045)** | **Profile 4**  **(n=1,074)** | **Profile 5**  **(n=4,158)** |
| --- | --- | --- | --- | --- | --- |
| Sex, male, n (%) | 1892(61.28%) | 34217(49.22%) | 1967(48.63%) | 848(78.96%) | 3007(72.32%) |
| Age end of follow-up, years, *m* (SD) | 13(1.96) | 13(1.14) | 13(1.95) | 13(1.74) | 13(1.80) |
| Emotional disorder, n(%) | 138(4.47%) | 1829 (2.63%) | 249 (6.16%) | 87 (8.10%) | 118 (2.84%) |
| Anxiety disorder, n(%) | 51 (1.65%) | 793 (1.14%) | 134 (3.31%) | 22 (2.05%) | 49 (1.18%) |
| Depressive disorder, n(%) | 17 (0.55%) | 387 (0.56%) | 26 (0.64%) | 8 (0.74%) | 19 (0.46%) |
| Childhood onset emotional disorder, n(%) | 33 (1.07%) | 178 (0.26%) | 62 (1.53%) | 38 (3.54%) | 18 (0.43%) |
| Adolescent onset emotional disorder, n(%) | 105 (3.40%) | 1651 (2.37%) | 187 (4.62%) | 49 (4.56%) | 100 (2.41%) |
| ADHD, n(%) | 575 (18.63%) | 1771 (2.55%) | 228 (5.64%) | 317 (29.52%) | 265 (6.37%) |
| ODD, n(%) | 54 (1.75%) | 243 (0.35%) | 25 (0.62%) | 61 (5.68%) | 42 (1.01%) |
| OCD,n(%) | 17 (0.55%) | 247 (0.36%) | 39 (0.96%) | 11 (1.02%) | 21 (0.51%) |
| Highly educated mother or father, n (%) | 1873 (66.39%) | 48975 (75.38%) | 2482 (68.06%) | 660 (66.20%) | 3031 (77.74%) |
| Mother’s country of birth |  |  |  |  |  |
| Norway, n(%) | 2779 (91.66%) | 63704 (93.33%) | 3488 (87.40%) | 981 (92.90%) | 3836 (93.84%) |
| Other high-income-country, n (%) | 139 (4.58%) | 3059 (4.48%) | 199 (4.99%) | 52 (4.92%) | 174 (4.26%) |
| Other GDB 7 super region country, n(%) | 114 (3.76%) | 1492 (2.19%) | 304 (7.62%) | 23 (2.18%) | 78 (1.91%) |

### **Table S10** Association between mental health traits in childhood and any childhood-onset adolescent persistent emotional disorder (10-18 years).

| **Emotional disorder in adolescence (10-18 years)** | | | | |
| --- | --- | --- | --- | --- |
| **Temperament and personality traits** | **OR^1^** | **95% CI^1^** | **p-value** | **q-value^2^** |
| Fussy temperament 6mths | 1.19 | 1.08, 1.31 | <0.001 | <0.001 |
| Positive temperament 6mths | 0.93 | 0.85, 1.01 | 0.081 | 0.2 |
| Emotionality 18mths | 1.16 | 1.03, 1.30 | 0.013 | 0.029 |
| Emotionality 3yrs | 1.43 | 1.25, 1.64 | <0.001 | <0.001 |
| Emotionality 5yrs | 1.81 | 1.53, 2.13 | <0.001 | <0.001 |
| Shyness 18mths | 1.02 | 0.91, 1.15 | 0.7 | 0.7 |
| Shyness 3yrs | 1.15 | 1.01, 1.30 | 0.033 | 0.069 |
| Shyness 5yrs | 1.39 | 1.19, 1.63 | <0.001 | <0.001 |
| Sociability 18mths | 0.95 | 0.84, 1.08 | 0.4 | 0.5 |
| Sociability 3yrs | 0.94 | 0.82, 1.08 | 0.4 | 0.5 |
| Sociability 5yrs | 0.83 | 0.70, 0.97 | 0.021 | 0.046 |
| Activity 18mths | 1.11 | 0.97, 1.27 | 0.14 | 0.3 |
| Activity 3yrs | 1.27 | 1.09, 1.47 | 0.002 | 0.005 |
| Activity 5yrs | 1.14 | 0.93, 1.38 | 0.2 | 0.3 |
| Neuroticism 8yrs | 2.85 | 2.50, 3.24 | <0.001 | <0.001 |
| Conscientiousness 8yrs | 0.62 | 0.53, 0.71 | <0.001 | <0.001 |
| Agreeableness 8yrs | 0.51 | 0.45, 0.59 | <0.001 | <0.001 |
| Imagination 8yrs | 0.62 | 0.54, 0.71 | <0.001 | <0.001 |
| Extraversion 8yrs | 0.63 | 0.54, 0.72 | <0.001 | <0.001 |
| **Mental health problems** |  |  |  |  |
| Emotional problems 18 mths | 1.26 | 1.14, 1.39 | <0.001 | <0.001 |
| Emotional problems 3yrs | 1.43 | 1.30, 1.57 | <0.001 | <0.001 |
| Emotional problems 5 yrs | 1.86 | 1.72, 2.00 | <0.001 | <0.001 |
| Behavioral problems 18 mths | 1.26 | 1.14, 1.41 | <0.001 | <0.001 |
| Behavioral problems 3yrs | 1.68 | 1.50, 1.89 | <0.001 | <0.001 |
| Behavioral problems 5 yrs | 1.84 | 1.63, 2.09 | <0.001 | <0.001 |
| Depressive symptoms 8yrs | 1.84 | 1.73, 1.96 | <0.001 | <0.001 |
| Anxiety symptoms 8yrs | 2.17 | 2.00, 2.35 | <0.001 | <0.001 |
| Inattention 8yrs | 1.78 | 1.61, 1.98 | <0.001 | <0.001 |
| Hyperactivity 8yrs | 1.79 | 1.63, 1.96 | <0.001 | <0.001 |
| Conduct disorder symptoms 8yrs | 1.57 | 1.46, 1.69 | <0.001 | <0.001 |
| Oppositional defiant disorder symptoms 8yrs | 1.96 | 1.81, 2.12 | <0.001 | <0.001 |
| ^1^OR = Odds Ratio per standard deviation change in trait score, CI = Confidence Interval | | | | |
| ^2^False discovery rate correction for multiple testing. Q-value calculated for all 31 evaluated measures (temperament traits and emotional symptoms). | | | | |
| OR for each scale is calculated separately in logistic regression with birth year and sex as covariates. Calculated using robust standard errors HC0 and clustered for maternal ID. | | | | |

### **Table S11** Association between developmental trajectories and any childhood-onset adolescent persistent emotional disorder

| **Comparison** | **Odds ratio (OR)** | **CI** |
| --- | --- | --- |
| Profile 1 vs Reference | 8.45 | (2.02, 14.89) |
| Profile 3 vs Reference | 14.81 | (6.45, 23.18) |
| Profile 4 vs Reference | 30.40 | (10.59, 50.22) |
| Profile 5 vs Reference | 3.38 | (0.68, 6.08) |
| Profile 1 vs profile 3 | 0.57 | (0.23, 0.92) |
| Profile 1 vs profile 4 | 0.28 | (0.09, 0.47) |
| Profile 1 vs profile 5 | 2.50 | (0.59, 4.41) |
| Profile 3 vs profile 4 | 0.49 | (0.25, 0.72) |
| Profile 3 vs profile 5 | 4.38 | (1.67, 7.09) |
| Profile 4 vs profile 5 | 9.00 | (3.14, 14.85) |

**Note:** Profile 2 including 84.9% (n=69522) of the individuals was used as reference class. Distribution of individuals to subsequent classes: Profile 1 (n=3087, 3.77%), Profile 3 (n=4045, 4.94%), Profile 4 (n=1074, 1.31%), Profile5 (n=4158, 5.01%). All profiles were compared to each other. All associations were significant with p<0.001.

### **Table S12:** Association between mental health traits in childhood and any adolescent-onset emotional disorder (10-18 years).

| **Emotional disorder in adolescence (10-18 years)** | | | | |
| --- | --- | --- | --- | --- |
| **Temperament and personality traits** | **OR^1^** | **95% CI^1^** | **p-value** | **q-value^2^** |
| Fussy temperament 6mths | 1.05 | 1.01, 1.10 | 0.019 | 0.025 |
| Positive temperament 6mths | 1.01 | 0.97, 1.05 | 0.7 | 0.7 |
| Emotionality 18mths | 1.07 | 1.02, 1.12 | 0.003 | 0.004 |
| Emotionality 3yrs | 1.15 | 1.09, 1.22 | <0.001 | <0.001 |
| Emotionality 5yrs | 1.29 | 1.19, 1.39 | <0.001 | <0.001 |
| Shyness 18mths | 1.00 | 0.95, 1.04 | 0.9 | >0.9 |
| Shyness 3yrs | 1.03 | 0.97, 1.09 | 0.3 | 0.4 |
| Shyness 5yrs | 1.08 | 0.99, 1.18 | 0.074 | 0.090 |
| Sociability 18mths | 0.97 | 0.93, 1.02 | 0.3 | 0.3 |
| Sociability 3yrs | 0.96 | 0.90, 1.01 | 0.11 | 0.13 |
| Sociability 5yrs | 0.97 | 0.89, 1.05 | 0.4 | 0.5 |
| Activity 18mths | 1.06 | 1.01, 1.11 | 0.027 | 0.033 |
| Activity 3yrs | 1.03 | 0.97, 1.08 | 0.4 | 0.4 |
| Activity 5yrs | 1.07 | 0.98, 1.17 | 0.11 | 0.13 |
| Neuroticism 8yrs | 1.63 | 1.52, 1.74 | <0.001 | <0.001 |
| Conscientiousness 8yrs | 0.83 | 0.78, 0.90 | <0.001 | <0.001 |
| Agreeableness 8yrs | 0.76 | 0.71, 0.82 | <0.001 | <0.001 |
| Imagination 8yrs | 0.86 | 0.80, 0.92 | <0.001 | <0.001 |
| Extraversion 8yrs | 0.90 | 0.84, 0.97 | 0.004 | 0.005 |
| **Mental health problems** |  |  |  |  |
| Emotional problems 18 mths | 1.14 | 1.08, 1.19 | <0.001 | <0.001 |
| Emotional problems 3yrs | 1.18 | 1.13, 1.24 | <0.001 | <0.001 |
| Emotional problems 5 yrs | 1.37 | 1.30, 1.46 | <0.001 | <0.001 |
| Behavioral problems 18 mths | 1.09 | 1.05, 1.14 | <0.001 | <0.001 |
| Behavioral problems 3yrs | 1.22 | 1.15, 1.28 | <0.001 | <0.001 |
| Behavioral problems 5 yrs | 1.34 | 1.25, 1.44 | <0.001 | <0.001 |
| Depressive symptoms 8yrs | 1.39 | 1.33, 1.45 | <0.001 | <0.001 |
| Anxiety symptoms 8yrs | 1.45 | 1.38, 1.53 | <0.001 | <0.001 |
| Inattention 8yrs | 1.28 | 1.22, 1.35 | <0.001 | <0.001 |
| Hyperactivity 8yrs | 1.25 | 1.18, 1.32 | <0.001 | <0.001 |
| Conduct disorder symptoms 8yrs | 1.21 | 1.15, 1.27 | <0.001 | <0.001 |
| Oppositional defiant disorder symptoms 8yrs | 1.38 | 1.31, 1.46 | <0.001 | <0.001 |
| ^1^OR = Odds Ratio per standard deviation change in trait score, CI = Confidence Interval | | | | |
| ^2^False discovery rate correction for multiple testing. Q-value calculated for all 31 evaluated measures (temperament traits and emotional symptoms). | | | | |
| OR for each scale is calculated separately in logistic regression with birth year and sex as covariates. Calculated using robust standard errors HC0 and clustered for maternal ID. | | | | |

### **Table S13** Association between developmental trajectories and any adolescent-onset emotional disorder

| **Comparison** | **Odds ratio (OR)** | **CI** |
| --- | --- | --- |
| Profile 1 vs Reference | 1.81 | (1.27, 2.35) |
| Profile 3 vs Reference | 2.75 | (2,00, 3.00) |
| Profile 4 vs Reference | 2.98 | (2.21, 3.30) |
| Profile 5 vs Reference | 1.28 | (0.96, 1.61) |
| Profile 1 vs profile 3 | 0.66 | (0.43, 0.88) |
| Profile 1 vs profile 4 | 0.61 | (0.33, 0.90) |
| Profile 1 vs profile 5 | 1.41 | (0.88, 1.94) |
| Profile 3 vs profile 4 | 0.93 | (0.58, 1.27) |
| Profile 3 vs profile 5 | 2.15 | (1.52, 2.77) |
| Profile 4 vs profile 5 | 2.31 | (1.40, 3.23) |

**Note:** Profile 2 including 84.9% (n=69522) of the individuals was used as reference class. Distribution of individuals to subsequent classes: Profile 1 (n=3087, 3.77%), Profile 3 (n=4045, 4.94%), Profile 4 (n=1074, 1.31%), Profile5 (n=4158, 5.01%). All profiles were compared to each other. All associations were significant with p<0.001.

### **Table S14** Association between mental health traits in childhood and any adolescent emotional disorder (10-18 years) (males only)

| **Emotional disorder in adolescence (10-18 years)** | | | | |
| --- | --- | --- | --- | --- |
| **Temperament and personality traits** | **OR^1^** | **95% CI^1^** | **p-value** | **q-value^2^** |
| Fussy temperament 6mths | 1.06 | 1.00, 1.12 | 0.035 | 0.039 |
| Positive temperament 6mths | 0.99 | 0.93, 1.04 | 0.6 | 0.7 |
| Emotionality 18mths | 1.09 | 1.03, 1.16 | 0.005 | 0.006 |
| Emotionality 3yrs | 1.25 | 1.16, 1.34 | <0.001 | <0.001 |
| Emotionality 5yrs | 1.46 | 1.32, 1.60 | <0.001 | <0.001 |
| Shyness 18mths | 0.98 | 0.92, 1.04 | 0.5 | 0.5 |
| Shyness 3yrs | 1.05 | 0.97, 1.13 | 0.2 | 0.2 |
| Shyness 5yrs | 1.15 | 1.04, 1.28 | 0.007 | 0.008 |
| Sociability 18mths | 0.99 | 0.93, 1.05 | 0.7 | 0.7 |
| Sociability 3yrs | 1.00 | 0.92, 1.08 | >0.9 | >0.9 |
| Sociability 5yrs | 0.93 | 0.84, 1.03 | 0.2 | 0.2 |
| Activity 18mths | 1.12 | 1.05, 1.20 | 0.001 | 0.001 |
| Activity 3yrs | 1.15 | 1.06, 1.24 | <0.001 | <0.001 |
| Activity 5yrs | 1.11 | 1.00, 1.23 | 0.061 | 0.067 |
| Neuroticism 8yrs | 1.89 | 1.74, 2.05 | <0.001 | <0.001 |
| Conscientiousness 8yrs | 0.76 | 0.69, 0.82 | <0.001 | <0.001 |
| Agreeableness 8yrs | 0.64 | 0.59, 0.70 | <0.001 | <0.001 |
| Imagination 8yrs | 0.80 | 0.73, 0.88 | <0.001 | <0.001 |
| Extraversion 8yrs | 0.81 | 0.74, 0.89 | <0.001 | <0.001 |
| **Mental health problems** |  |  |  |  |
| Emotional problems 18 mths | 1.17 | 1.10, 1.24 | <0.001 | <0.001 |
| Emotional problems 3yrs | 1.24 | 1.16, 1.33 | <0.001 | <0.001 |
| Emotional problems 5 yrs | 1.52 | 1.42, 1.62 | <0.001 | <0.001 |
| Behavioral problems 18 mths | 1.16 | 1.10, 1.23 | <0.001 | <0.001 |
| Behavioral problems 3yrs | 1.37 | 1.28, 1.46 | <0.001 | <0.001 |
| Behavioral problems 5 yrs | 1.49 | 1.38, 1.61 | <0.001 | <0.001 |
| Depressive symptoms 8yrs | 1.57 | 1.49, 1.65 | <0.001 | <0.001 |
| Anxiety symptoms 8yrs | 1.68 | 1.57, 1.79 | <0.001 | <0.001 |
| Inattention 8yrs | 1.37 | 1.29, 1.46 | <0.001 | <0.001 |
| Hyperactivity 8yrs | 1.38 | 1.30, 1.47 | <0.001 | <0.001 |
| Conduct disorder symptoms 8yrs | 1.31 | 1.24, 1.37 | <0.001 | <0.001 |
| Oppositional defiant disorder symptoms 8yrs | 1.57 | 1.47, 1.67 | <0.001 | <0.001 |
| ^1^OR = Odds Ratio per standard deviation change in trait score, CI = Confidence Interval | | | | |
| ^2^False discovery rate correction for multiple testing. Q-value calculated for all 31 evaluated measures (temperament traits and emotional symptoms). | | | | |
| OR for each scale is calculated separately in logistic regression with birth year and sex as covariates. Calculated using robust standard errors HC0 and clustered for maternal ID. | | | | |

### **Table S15** Association between developmental trajectories and any adolescent emotional disorder (males only)

| **Comparison** | **Odds ratio (OR)** | **CI** |
| --- | --- | --- |
| Profile 1 vs Reference | 2.13 | (1.26,3.01) |
| Profile 3 vs Reference | 4.92 | (3.70,6.15) |
| Profile 4 vs Reference | 4.90 | (3.25,6.56) |
| Profile 5 vs Reference | 1.78 | (1.27,2.30) |
| Profile 3 vs profile 1 | 2.31 | (1.30,3.32) |
| Profile 3 vs profile 4 | 1.00 | (0.62,1.39) |
| Profile 3 vs profile 5 | 2.76 | (1.89,3.64) |
| Profile 5 vs profile 1 | 0.84 | (0.45,1.22) |
| Profile 5 vs profile 4 | 0.36 | (0.34,0.40) |
| Profile 1 vs profile 4 | 0.44 | (0.39,0.46) |

**Note:** Profile 2 including 83.8% (n=35120) of the individuals was used as reference class. Distribution of individuals to subsequent classes: Profile1 (n=1562, 3.73%), Profile 3 (n=1875, 4.72%), Profile 4 (n=788, 1.88%), Profile 5 (n=2586, 6.17%).Number of individuals diagnosed with emotional disorder in each profile: Profile1(n=60, 3.84%), Profile 2(n=819, 2.33%), Profile 3 (n=131,7.0%), Profile4 (n=58, 7.36%), Profile5 (n=84,3.25%). All profiles were compared to each other. All associations were significant with p<0.001.

### **Table S16** Association between mental health traits in childhood and any adolescent emotional disorder (10-18 years) (females only)

| **Emotional disorder in adolescence (10-18 years)** | | | | |
| --- | --- | --- | --- | --- |
| **Temperament and personality traits** | **OR^1^** | **95% CI^1^** | **p-value** | **q-value^2^** |
| Fussy temperament 6mths | 1.08 | 1.02, 1.14 | 0.004 | 0.006 |
| Positive temperament 6mths | 1.00 | 0.95, 1.06 | 0.9 | 0.9 |
| Emotionality 18mths | 1.08 | 1.02, 1.14 | 0.014 | 0.017 |
| Emotionality 3yrs | 1.15 | 1.07, 1.23 | <0.001 | <0.001 |
| Emotionality 5yrs | 1.29 | 1.16, 1.44 | <0.001 | <0.001 |
| Shyness 18mths | 1.02 | 0.96, 1.08 | 0.5 | 0.5 |
| Shyness 3yrs | 1.04 | 0.97, 1.12 | 0.2 | 0.3 |
| Shyness 5yrs | 1.12 | 1.00, 1.26 | 0.043 | 0.049 |
| Sociability 18mths | 0.95 | 0.90, 1.01 | 0.11 | 0.13 |
| Sociability 3yrs | 0.91 | 0.85, 0.98 | 0.013 | 0.016 |
| Sociability 5yrs | 0.95 | 0.85, 1.06 | 0.3 | 0.4 |
| Activity 18mths | 1.02 | 0.96, 1.08 | 0.6 | 0.6 |
| Activity 3yrs | 0.98 | 0.91, 1.05 | 0.6 | 0.6 |
| Activity 5yrs | 1.05 | 0.94, 1.19 | 0.4 | 0.4 |
| Neuroticism 8yrs | 1.73 | 1.58, 1.89 | <0.001 | <0.001 |
| Conscientiousness 8yrs | 0.82 | 0.75, 0.91 | <0.001 | <0.001 |
| Agreeableness 8yrs | 0.78 | 0.72, 0.86 | <0.001 | <0.001 |
| Imagination 8yrs | 0.81 | 0.74, 0.89 | <0.001 | <0.001 |
| Extraversion 8yrs | 0.87 | 0.80, 0.96 | 0.004 | 0.005 |
| **Mental health problems** |  |  |  |  |
| Emotional problems 18 mths | 1.14 | 1.07, 1.22 | <0.001 | <0.001 |
| Emotional problems 3yrs | 1.21 | 1.14, 1.28 | <0.001 | <0.001 |
| Emotional problems 5 yrs | 1.47 | 1.37, 1.58 | <0.001 | <0.001 |
| Behavioral problems 18 mths | 1.07 | 1.01, 1.14 | 0.021 | 0.025 |
| Behavioral problems 3yrs | 1.20 | 1.12, 1.29 | <0.001 | <0.001 |
| Behavioral problems 5 yrs | 1.38 | 1.25, 1.53 | <0.001 | <0.001 |
| Depressive symptoms 8yrs | 1.45 | 1.37, 1.55 | <0.001 | <0.001 |
| Anxiety symptoms 8yrs | 1.57 | 1.46, 1.68 | <0.001 | <0.001 |
| Inattention 8yrs | 1.41 | 1.30, 1.52 | <0.001 | <0.001 |
| Hyperactivity 8yrs | 1.35 | 1.24, 1.47 | <0.001 | <0.001 |
| Conduct disorder symptoms 8yrs | 1.28 | 1.18, 1.40 | <0.001 | <0.001 |
| Oppositional defiant disorder symptoms 8yrs | 1.44 | 1.34, 1.55 | <0.001 | <0.001 |
| ^1^OR = Odds Ratio per standard deviation change in trait score, CI = Confidence Interval | | | | |
| ^2^False discovery rate correction for multiple testing. Q-value calculated for all 31 evaluated measures (temperament traits and emotional symptoms). | | | | |
| OR for each scale is calculated separately in logistic regression with birth year and sex as covariates. Calculated using robust standard errors HC0 and clustered for maternal ID. | | | | |

### **Table S17** Association between developmental trajectories and any adolescent emotional disorder (females only).

| **Comparison** | **Odds ratio (OR)** | **CI** |
| --- | --- | --- |
| Profile 1 vs Reference | 2.44 | (1.60,3.28) |
| Profile 3 vs Reference | 2.86 | (2.18,3.56) |
| Profile 4 vs Reference | 3.46 | (1.94,4.98) |
| Profile 5 vs Reference | 1.07 | (0.73,1.40) |
| Profile 3 vs profile 1 | 1.18 | (0.71,1.64) |
| Profile 3 vs profile 4 | 0.83 | (0.43,1.23) |
| Profile 3 vs profile 5 | 2.68 | (1.71,3.66) |
| Profile 5 vs profile 1 | 0.44 | (0.24,0.63) |
| Profile 5 vs profile 4 | 0.31 | (0.15,0.48) |
| Profile 1 vs profile 4 | 0.70 | (0.31,1.10) |

**Note:** Profile 2 including 85.4% (n=34139) of the individuals was used as reference class. Distribution of individuals to subsequent classes: Profile 1 (n=1360, 3.40%), Profile 3 (n=2195, 5.49%), Profile4 (n=318, 0.80%), Profile 5 (n=1943, 4.86%). Number of individuals diagnosed with emotional disorder in each profile: Profile1(n=69, 5.07%), Profile 2(n=998, 2.92%), Profile 3 (n=126, 5.74%), Profile4 (n=25, 7.86%), Profile5 (n=51,2.62%). All profiles were compared to each other. All associations were significant with p<0.001.
